# Single cell AI-based detection of DNA mismatch repair deficiency in 1,988 colorectal cancers reveals prognostic and predictive value in the SCOT trial

**DOI:** 10.1101/2023.12.18.23300137

**Authors:** Marta Nowak, Faiz Jabbar, Ann-Katrin Rodewald, Luciana Gneo, Tijana Tomasevic, Andrea Harkin, Tim Iveson, Mark Saunders, Rachel Kerr, Karin Oein, Noori Maka, Jennifer Hay, Joanne Edwards, Ian Tomlinson, Owen Sansom, Caroline Kelly, Alistair Easton, Enric Domingo, TransSCOT group, Viktor H Koelzer, David N Church

## Abstract

Testing for DNA mismatch repair deficiency (MMRd) is recommended for all colorectal cancers (CRC). Automation of this would facilitate precision medicine, particularly if it provided information on likely aetiology. We developed AIMMer, an **AI**-based method for determination of **MMR** protein expression at single cell level in routine pathology samples. We applied it to over 2,000 colorectal cancers (CRC) from the SCOT clinical trial, which compared 3 vs 6 months of oxaliplatin-based adjuvant chemotherapy (FOLFOX or CAPOX). Benchmarking of AIMMeR against pathologist ground truth MMR calls revealed AUROC of 0.98, and positive predictive value (PPV) greater than 95% for the commonest pa[ern of somatic MMRd, and for retained MMR expression. Analysis of CRC recurrence confirmed the prognostic value of MMRd in oxaliplatin-treated patients. While MMRd did not predict differential benefit from chemotherapy duration, it correlated with difference in clinical outcome by chemotherapy regimen (*P*_Interaction_=0.04). AIMMeR holds promise to reduce pathologist workflow and streamline clinical diagnostics in CRC.

## INTRODUCTION

Colorectal cancer (CRC) is the third most common tumour globally, and a substantial cause of morbidity and mortality^1^. 10-15% of CRCs display genomic instability due to DNA mismatch repair deficiency (MMRd), caused by either germline mutation of MMR genes *MLH1*, *MSH2*, *MSH6* or *PMS2* (Lynch syndrome)^2^, biallelic somatic mutation of MMR genes^3,4^, or more commonly, somatic silencing of *MLH1* by promoter methylation^5^. MMRd causes failure to repair errors accumulated during DNA replication – particularly those at error-prone DNA microsatellites – resulting in elevated tumour mutational burden (TMB) and microsatellite instability (MSI). MMRd CRC display characteristic clinical and pathological features, including right-sided colonic location, female preponderance, prominent lymphocytic infiltrate and good prognosis in early-stage disease^6–9^. They are also highly sensitive to immune checkpoint blockade (ICB), resulting in prolonged disease control in the metastatic setting^10,11^, and unprecedented pathological responses in localised disease that have raised the possibility of organ-sparing therapy^12,13^. In view of these important correlates, reflex immunohistochemistry (IHC) for MMRd or MSI PCR is recommended for all incident CRCs by international guidelines^14–16^. In most centres, MMR IHC is preferred, with slides or images reviewed by specialised GI pathologists and MMRd identified by loss of MMR protein expression in tumour epithelium. This requires substantial pathologist time. Consequently, efforts have focused on the development of automated methods to identify MMRd, leveraging advances in artificial intelligence (AI) and machine learning (ML) for image analysis. By applying AI-based deep learning to haematoxylin and eosin (H&E) stained slides, Kather and colleagues developed a method to identify MMRd in gastrointestinal cancers^17^, and refined this in a larger series of CRCs to give an AUROC ranging from 0.74 to 0.96^18,19^. While impressive, when used as a rule-out for additional MMR/MSI testing with fixed 95% sensitivity, this method still requires MMR testing with pathologist review in 23.2 to 87.4% cases, depending on the cohort analysed^19^. Furthermore, it provides no information about the likely aetiology of MMRd, detail regarding which is helpful in stratifying cases for germline testing^20,21^.

One possible alternative to H&E slides would be to use IHC-stained images of tumour tissue as the substrate for automated image analysis. Multiple studies, including from our group, have published methods for quantification of IHC labelled cells in whole tumours^22^ or defined intratumoral regions such as the tumour invasive margin^23^, intraepithelial or intrastromal compartments^24^. While defining MMR protein expression in cells within the tumour intraepithelial compartment is superficially attractive for detection of MMRd, this approach is undermined by the detection of intra-epithelial lymphocytes, which retain MMR protein expression and are enriched in MMRd tumours^22,25,26^. This shortcoming may be circumvented by the classification of MMR protein expression in individual cells, however methods to do this have previously been lacking. We sought to address this by developing a method for analysis of MMR status at the single cell level. By applying this to more than 2,000 CRCs from the SCOT clinical trial^27^, were able to benchmark our assay against the gold-standard of pathologist review, and to determine the clinical correlates of MMRd in this practice-changing study.

## RESULTS

### Development of AIMMeR: an AI-based methodology for single cell analysis of DNA mismatch repair loss

To develop and test AIMMeR – an AI-based method for detection of MMRd, we performed immunostaining for MLH1, MSH2, MSH6 and PMS2 on tissue microarrays (TMA) of tumors from the SCOT trial, which compared efficacy and tolerability of 6 vs 3 months of adjuvant oxaliplatin-based chemotherapy in patients with stage III and high-risk stage II CRC recruited from >100 sites^27^ (study CONSORT diagram provided as Figure 1). To minimise the risk of incorrect assignation of MMR status from misclassification of MMR protein-expressing non-epithelial cells within the intraepithelial compartment (e.g., lymphocytes), we developed a novel method to classify cells by their nuclear morphology (epithelial, stromal, lymphocyte) using the nuclear segmentation model and nuclear classifier of the HALO AI digital analysis software v3.3 (Methods, Figure S1). This method also identified additional relevant features, including non-nuclear objects such as apoptotic bodies or extracellular matrix components, and artefacts such as tissue folds and background staining (Figure S1). Comparison of single-cell type classification with expert pathologist ground truth demonstrated overall accuracy of 0.92 (Methods, Table S1). We combined the nuclear morphological analysis with automated identification of 3-3’Diaminobenzidine (DAB) positivity^24^, and used the combined method – AIMMeR, to determine individual MMR protein expression (present vs absent) in single cells, and thus, the percentage of epithelial and stromal cells expressing individual MMR proteins in TMA cores and cases (Figure 2). After removal of cases with duplicate or non-matching trial ID, and pre-analytic fails, we performed additional QC to exclude cases with insufficient number of tumour cells, failed staining, or those lacking successful staining for all four MMR proteins. Pathologist review of tumours with possible MMR loss (see below) resulted in exclusion of a further 27 cases (predominantly for inadequate immunostaining). Our dataset for analysis of AIMMeR performance included 2,015 tumors and 38,113,216 single cells.

**Figure 1.**
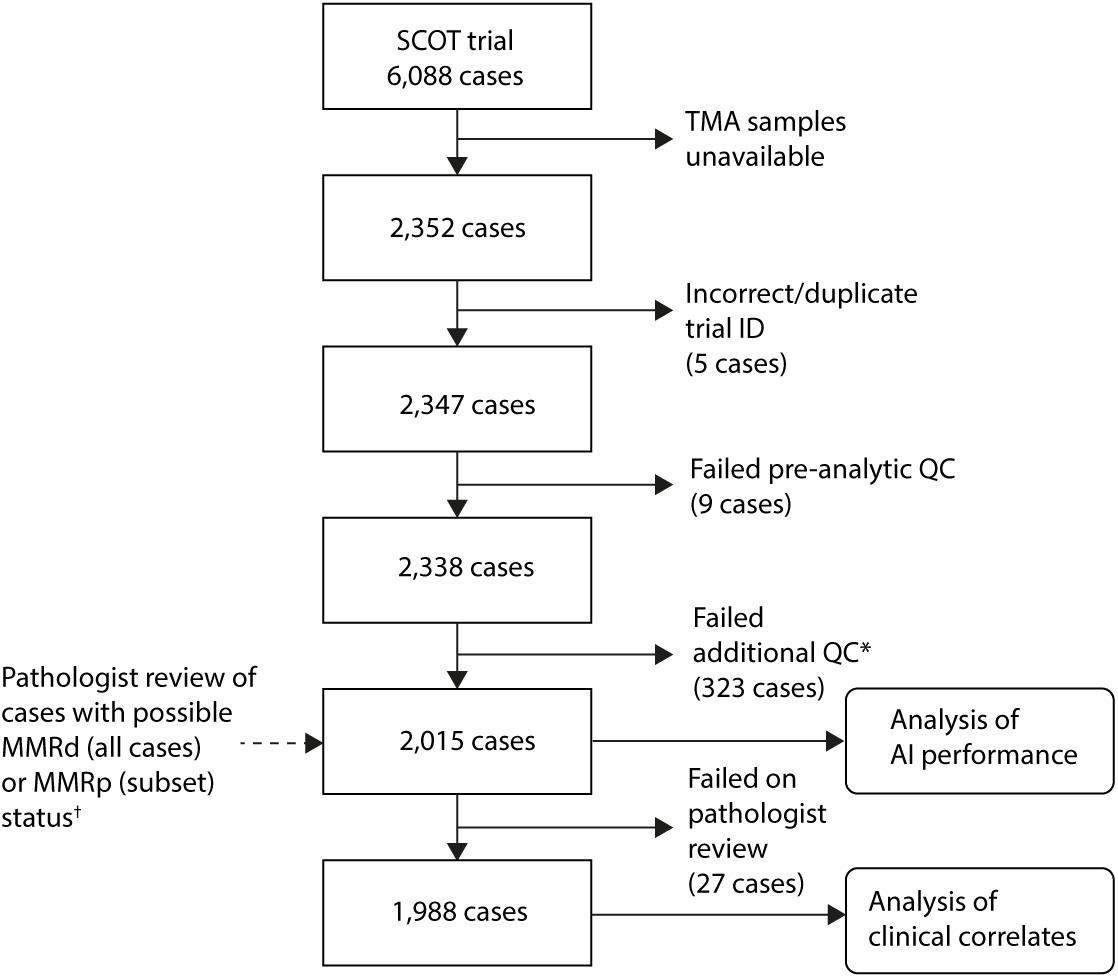
Flow (CONSORT) diagram of cases included in this study. *Additional QC included exclusion of tissue microarray (TMA) cores with <100 epithelial cells per core, cores with negative staining (<20 positive cells) for all four MMR proteins in both epithelium and stroma, and cases uninformative for all four MMR proteins. ^†^Pathologist review was performed in all 487 cases in which ≥1 core had <20% epithelial cells positive for ≥ 1 MMR protein expression, and a further 198 cases selected at random from 1,330 tumors in which all four MMR proteins were expressed in ≥20% epithelial cells in all TMA cores.

**Figure 2.**
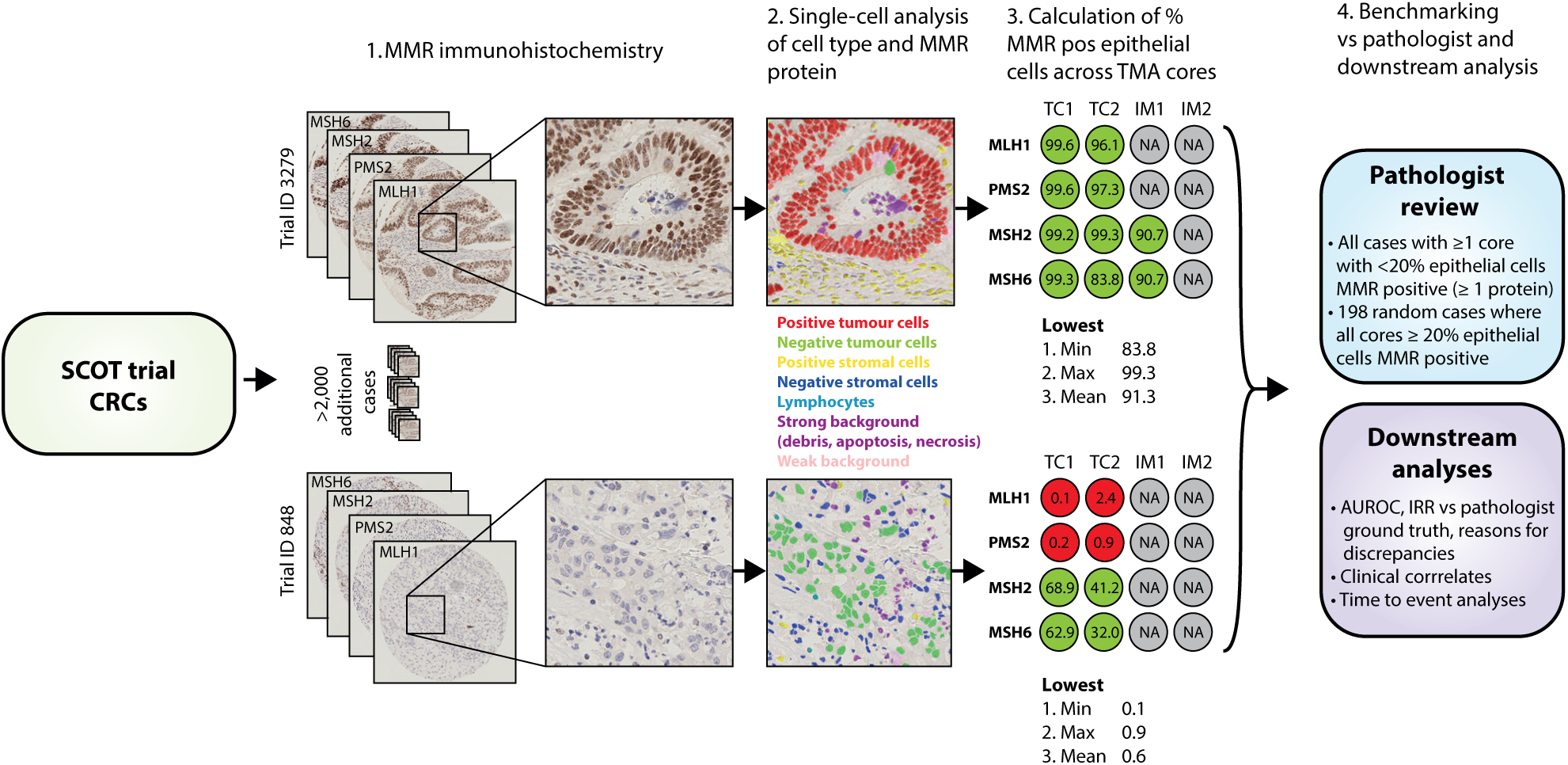
Schematic of study methodology. Immunohistochemistry (IHC) for DNA mismatch repair (MMR) proteins MLH1, PMS2, MSH2 and MSH6 was performed on tissue microarray (TMA) cores from 2,352 SCOT cases. For each case, one to four cores were available from the tumor center (TC1-4) and one to four from the invasive margin (IM1-4). Stained slides were scanned to generate digitized images, and cells within each core classified by AIMMeR according to type and MMR staining (see Methods and Results). Following QC, the percentage of epithelial cells positive for expression of individual MMR proteins was calculated for each core, and summary metrics were calculated for each case (shown in step 3). Cases with possible loss of MMR protein expression based on < 20% epithelial cells positive for ≥1 MMR protein in ≥1 TMA core (representative case ID 848 shown in lower panel) were identified and subject to expert pathologist review, together with a further 198 cases selected at random from 1,330 tumours in which all cores had ≥20% cells positive for all four MMR proteins (representative case ID 3279 shown in upper panel). Consensus pathologist ground truth was then established and used for benchmarking of AI performance and for analysis of clinical correlates of MMR loss. Further details are provided in the methods and the main text. Min – minimum; Max – maximum; AUROC – area under the receiver-operator curve; IRR – inter-rater reliability.

Initial analysis of AIMMeR results revealed anticipated variation in the percentage of epithelial cells expressing individual MMR proteins across cases (Figure 3a-e). Roughly two-thirds of tumours displayed homogenous positive staining for all four proteins (≥90% epithelial cells positive in all TMA cores), while ∼10% cases showed low or absent epithelial staining (<10% epithelial cells positive), most commonly in the case of MLH1 and PMS2 (Figure 3a). We found a strong positive correlation between cases in the percentage of epithelial cells positive for MLH1 and for PMS2 (Pearson r = 0.88, Spearman ρ = 0.79, both *P*<2.2e-16) and for MSH2 and MSH6 (Pearson r = 0.68, Spearman ρ = 0.74, both *P*<2.2e-16) (Figure 3f-h), consistent with MLH1-PMS2 and MSH2-MSH6 heterodimerisation; correlations for epithelial positivity of other MMR protein combinations were considerably less strong (Figure 3h, Figure S2). Interestingly, we also found strong positive correlation between cases in stromal expression of MMR proteins (Pearson r = 0.68–0.76, Spearman ρ = 0.70–0.78; *P*<2.2e-16 all cases), raising the possibility of coordinate regulation (Figure 3h, Figure S3).

**Figure 3.**
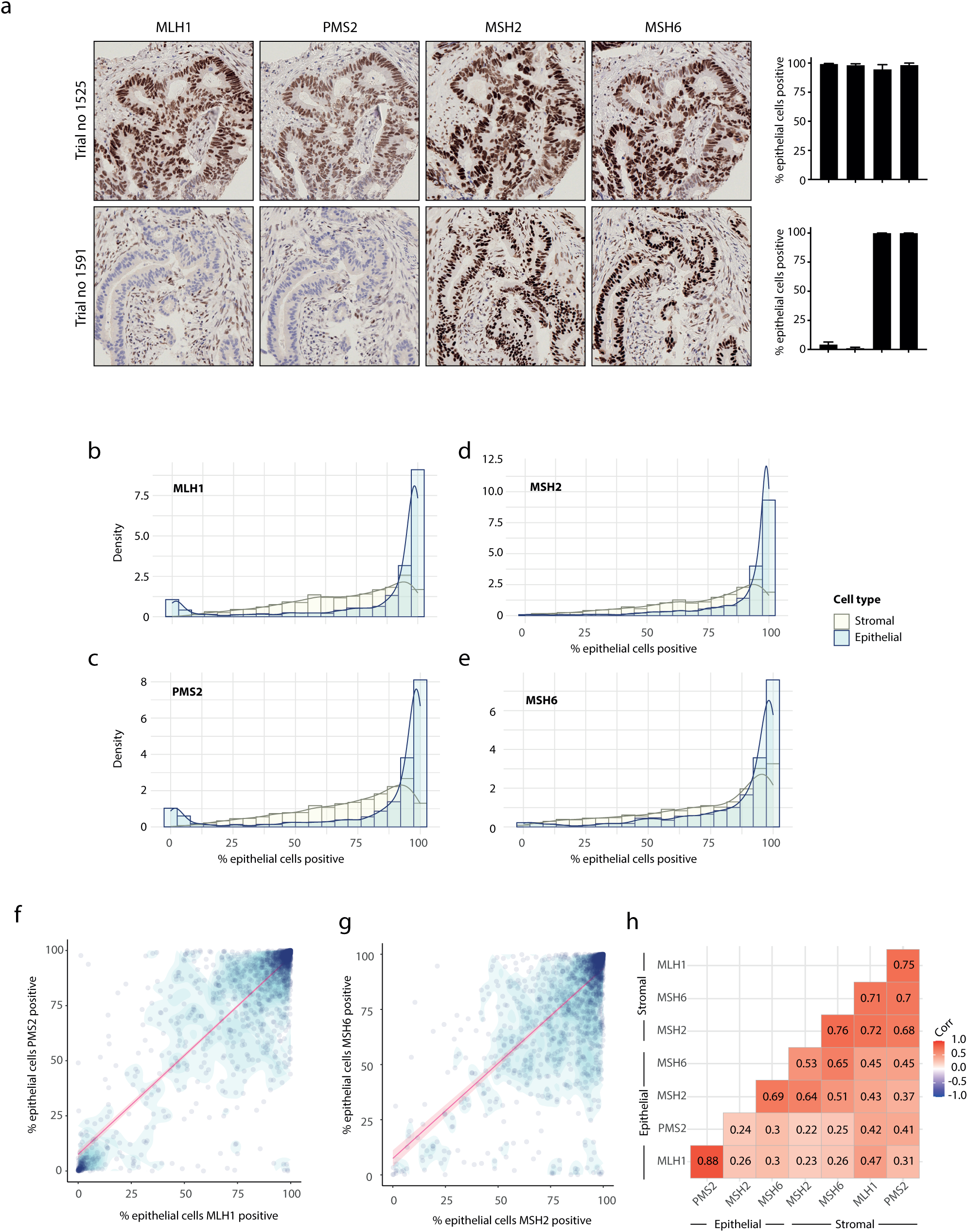
AIMMeR single cell analysis of MMR proteins identifies loss of epithelial expression and expected correlations. (a) Representative images of immunohistochemistry (IHC) for DNA mismatch repair (MMR) proteins MLH1, PMS2, MSH2 and MSH6 in cases with expression of all proteins (upper row), and loss of MLH1 and PMS2 with retained expression of MSH2 and MSH6 (lower row). Barplots to the right show the percentage of epithelial cells positive for each MMR protein as determined by AI (error bars indicate standard deviation between cores). (b-d) Frequency histograms with overlaid kernel density plots showing proportion of cases by percentage of epithelial and stromal cells positive for MMR proteins MLH1 (b), PMS2 (c), MSH2 (d) and MSH6 (e). (f, g) Scatterplots showing relationship between percentage of epithelial cells expressing dimerization partners MLH1 and PMS2 (f) and MSH2 and MSH6 (g). Regression line represents Spearman rho with 95% confidence intervals. (h) Matrix showing correlation between epithelial and stromal cell positivity for all four MMR proteins (P<2.2e-16 all cases). For consistency, plots in (b-h) show results from analysis of 1,988 cases and exclude 27 cases classed as failed on pathologist review; plots from analysis of original 2,015 cases were essentially identical.

### Identification of DNA mismatch repair deficiency (MMRd) by AIMMeR versus pathologist ground truth

We examined the discriminative ability of AIMMeR for detection of MMRd. Reasoning that most or all MMRd cases would have <20% epithelial cells positive for ≥1 MMR protein, and that tumors with ≥20% positive immunostaining for all MMR proteins would be MMRp, we selected all 487 cases with <20% immunostaining for any MMR protein, and 198 cases chosen at random from the >1,500 tumors with ≥20% immunostaining for all MMR proteins (Figure S4a). Scanned slides for these 685 cases were independently reviewed by two expert CRC pathologists (AE and VK), blinded to both the results of AIMMeR and to each other to generate individual pathologist MMR calls. Between-pathologist discrepancies were then resolved by discussion to generate consensus pathologist MMR calls. This consensus review identified 229 MMRd^28^ cases, all of which had <20% epithelial MMR immunostaining by AIMMeR. Every case with ≥20% positive immunostaining by AIMMeR was confirmed as MMRp. 27 cases were classified as failed owing to inadequate immunostaining – all of these had <15% positive epithelial staining for ≥1 MMR protein by AIMMeR (25 had <10% positive staining). Using these consensus calls as the ground truth, we calculated area under the receiver-operator curves (AUROCs) with internal validation by bootstrap to identify the lowest of the four mean (of TMA cores) percentages of positive epithelial cells across MMR proteins (i.e. the value from the MMR protein with the fewest positive epithelial cells) as the optimum predictor of MMRd (Figure S4a,b). This had an AUROC of 0.98 (bootstrap 95% CI = 0.97–0.99) (Figure 4a, Figure S4b) (corresponding out of bag estimate = 0.98; 95% CI = 0.97–0.99), and maximal Youden index (sensitivity plus specificity) of 1.87 at a cutpoint of 10.7% positive epithelial cells (Figure 4b). Calculation of inter-rater reliability in classification of MMR status between pathologist review (individual and consensus) and AI calls using this cutpoint revealed substantial or greater agreement (κ = 0.79–0.82; Gwet AC1 = 0.85–0.89), which approached the level observed between pathologists (κ = 0.88, Gwet AC1 = 0.92) as evidenced by overlapping 95% confidence intervals (Figure 4c, Table S2) (note that dependency of κ and AC1 on the prevalence of the categories under study means that these values are lower than anticipated from analysis of the whole cohort, as shown in Table S2). Of 262 cases classified as MMRd by AIMMeR, 212 (80.9%) were confirmed on consensus review, while 25 were reclassified as MMRp and 25 as failed (Figure 4d). For the 423 cases classified as MMRp by AIMMeR, 404 were confirmed as MMRp, while 17 were reclassified as MMRd and 2 as failed (Figure 4d). Thus, the positive predictive value (PPV) of AIMMeR for MMRd was 80.1%, while the negative predictive value (NPV) was 95.6%. While MMRd holds prognostic and predictive value in CRC, the pattern of protein loss permits triage of patients for further Lynch syndrome testing^20,21^. Although classification by AIMMeR again showed substantial agreement with individual and consensus pathologist calls, this was lower (κ = 0.66–0.69; Gwet AC1 = 0.79–0.82) and less than that between pathologists (κ = 0.84, Gwet AC1 = 0.91) (Figure 4c, Table S2). We noted substantial variation in the predictive performance of AIMMeR depending on the pattern of protein loss detected (Figure 4e, Table S4, Figure S5). For cases where AIMMeR reported combined MLH1-PMS2 loss, or combined MSH2-MSH6 loss, the PPV for MMRd was 96.6% and the PPV for the same combination of protein loss was 95.2%. In contrast, single MMR protein loss identified by AIMMeR had PPV of 61.8% for MMRd and 21.3% for single protein loss on consensus review, and other patterns of loss had corresponding PPVs of 57.7% and 11.5% respectively. The PPV of retained expression by AIMMeR classification was 95.5% for MMRp status (Figure 4e, Table S3, Figure S5). We analysed the reasons for discrepancies between AIMMeR and pathologist consensus MMR and protein calls. By far the most common was poor quality immunostaining, with failed, weak or background staining collectively accounting for 89.9% of discordances in MMR status, and 74.0% of discordances in protein expression (in cases where MMR status was concordant) (Figure 4f, Table S4, Figure S6). Interestingly, these reasons also accounted for most discrepancies in MMR and protein calls between pathologists (Figure 4f, Table S5). Other causes of discordance included technical issues (e.g., tissue folding) and rarely, atypical epithelial morphology (Figure 4f, Table S4, Figure S6).

**Figure 4.**
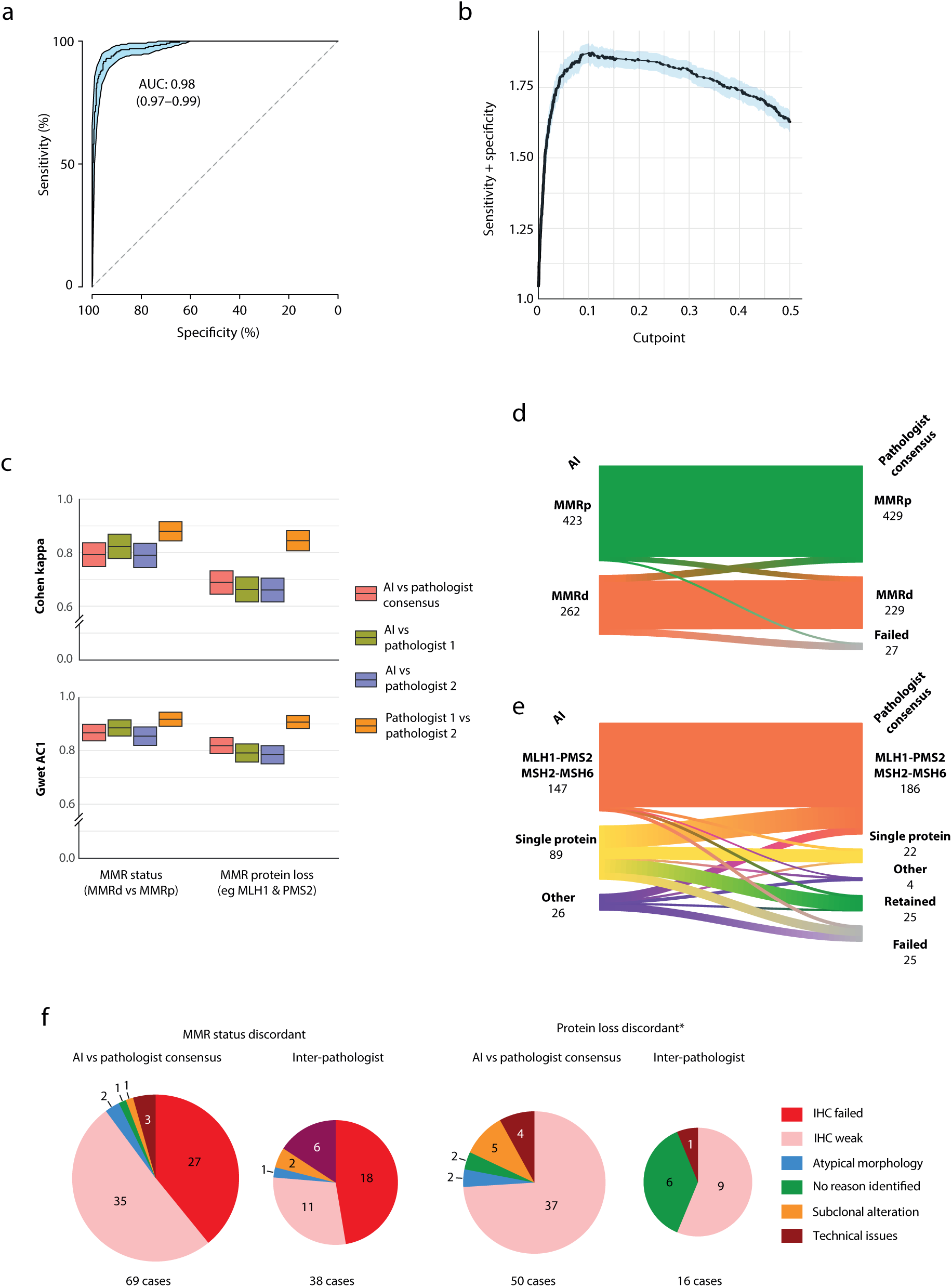
AIMMeR identifies mismatch repair deficiency with performance comparable to expert pathologists. (a) Area under the receiver-operator curve (AUROC) curve for AI-based identification of MMRd vs consensus pathologist ground truth in 685 cases (see text for details of selection). 95% confidence intervals were obtained by bootstrap (1,000 runs). (b) Optimum cut point for identification of MMRd by AI determined by bootstrap (1,000 runs) in 685 cases. (c) Inter-rater reliability measures of agreement between AI-based classification of tumour MMR status (left) and combination of MMR protein loss (right) vs individual and consensus pathologist classification. Measures of between-pathologist agreement are shown for comparison. AI-based classification uses a cut point of <10.7% positive epithelial cells in ≥1 MMR protein to define MMRd and individual protein loss. (d) Sankey plot showing relationship between MMR status determined by AI and following consensus pathologist review. (e) Sankey plot showing relationship between classification of MMR protein loss by AI and following consensus pathologist review. Category of “other” includes MLH1-PMS2 or MSH2-MSH6 loss plus other MMR proteins as well as alternative combinations of loss. (f) Reasons for discordance between AI and consensus pathologist calls identified at discrepancy review for both MMR status and protein status (*limited to cases for which MMR status was concordant). Reasons for discordance between pathologists are provided for comparison. Additional detail is provided in Tables S4,S5; illustrative cases are shown in Figure S6.

### Combined AIMMeR and pathologist classification of MMRd shows prognostic and predictive in SCOT trial cohort

We combined the consensus pathologist calls for the 658 tumours which passed review with the AIMMeR calls for the remaining 1,330 cases to determine the correlates of MMRd in the SCOT trial population. As expected, MMRd tumours were associated with older age, female sex and right-sided location (all *P*<0.0001) (Table S6). They also displayed significantly denser lymphocytic infiltrate, and significantly higher tumour/stromal ratio as determined by AI-based analysis (Figure S7a, b). Consistent with previous literature, patients with MMRd tumors had longer recurrence-free interval (RFI) (univariable hazard ratio [HR] = 0.69, 95% CI=0.50–0.96, *P*=0.027; multivariable-adjusted HR = 0.62, 95% CI = 0.44–0.88, *P*=0.007) (Figure 5a, Table S7). Analysis in prespecified subgroups suggested the prognostic value of MMRd was independent of patient sex and disease stage; concordant with prior results, the effect size was greater in younger patients (<70 years) and right-sided tumours, although interaction tests were not significant (Figure 5b). We found no evidence that MMRd predicted differential benefit from duration of chemotherapy (Figure 5c). Analysis by chemotherapy regimen revealed patients with MMRp tumors treated with CAPOX had similar rate of recurrence to those who had FOLFOX (univariable HR = 1.05, 95% CI = 0.86-1.29, *P*= 0.62; multivariable HR = 0.95, 95% CI= 0.77-1.16, *P*=0.6), while in contrast patients with MMRd tumours treated with FOLFOX had significantly worse outcomes (univariable HR = 1.90, 95% CI = 1.01-3.57, *P*= 0.046; multivariable HR = 2.08, 95% CI= 1.09-3.97, *P*=0.027, *P*_INTERACTION_=0.04). Although intriguing, this result should be interpreted with caution as chemotherapy regimen was chosen by the treating physician, rather than by randomisation.

**Figure 5.**
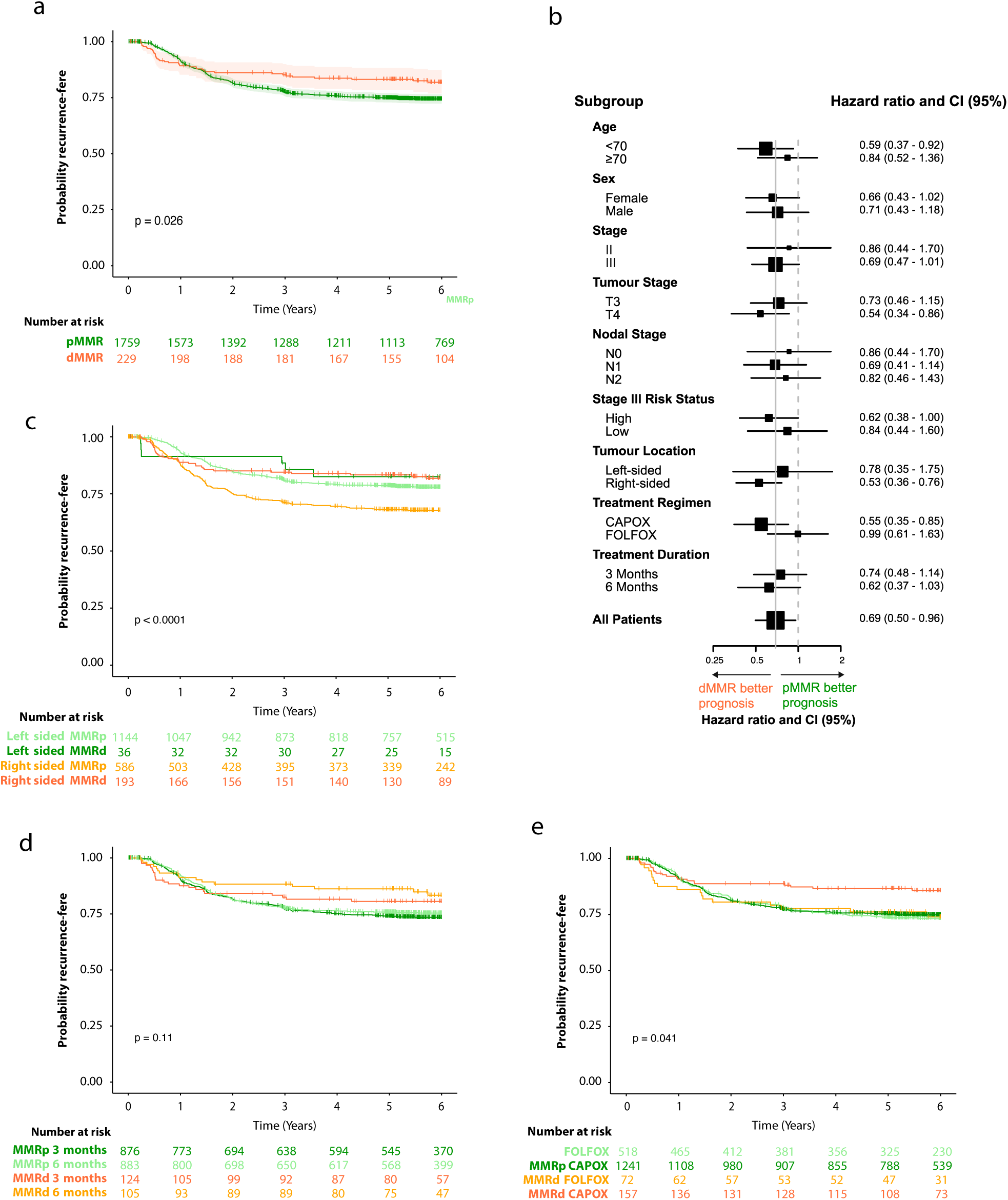
Prognostic and predictive value of combined AIMMeR and pathologist classification of MMRd in SCOT trial cohort. (a) Kaplan-Meier plot showing recurrence-free interval (RFI) for patients according to MMR status. (b) Forest plot showing hazard ratios (HR) with 95% confidence intervals (95% CI) for RFI according to MMR status within clinical and pathological subgroups by multivariable analysis*. (c) Kaplan-Meier plot showing RFI according to duration of chemotherapy and MMR status. (d) Kaplan-Meir plot showing RFI according to chemotherapy regimen and MMR status. HR in (a,c,d) were obtained by multivariable analysis*. * Multivariable Cox proportional hazards model included prespecified covariables of age, sex, pN stage, pT stage, sidedness, treatment regimen, treatment duration.

## DISCUSSION

To the best of our knowledge, we present the first study to identify loss of DNA mismatch repair in cancer at the single cell level. Our method, AIMMeR, was able to identify MMRd in CRC with performance approaching that of expert human pathologists. While its ability to determine the precise pattern of protein loss was lower, likely owing to the greater challenge this problem poses, AI-based identification of the most common combinations of protein loss – MLH1 and PMS2 or MSH2 and MSH6 – had PPV of greater than 95% for the same combination on consensus pathologist review. Importantly, AIMMeR classification of cases as MMRp had PPV of 95.5% for MMRp status on consensus pathologist review for all cases that underwent this, and 100% for the subset with ≥20% positive epithelial cells for all proteins. While additional testing and validation is required, AIMMeR holds promise for clinical application as a tool to reduce pathologist workload and streamline diagnostic workflows.

As noted above, previous efforts to identify MMRd in CRC by AI have largely focused on the use of H&E slide images, as these are inexpensive and are routinely generated as part of histopathological diagnosis^17–19,29^. Deep learning methods using whole slide images have been proposed as ‘rule out tests’ in CRC to remove the requirement for MMR immunostaining with pathologist review or MSI testing in cases classified as likely MMRp^19^. However, using current state of the art methods, on average roughly half of all tumours require additional testing with pathologist review^18,19^. Although AIMMeR requires MMR immunostaining of all tumours, this is both simple and routinely performed in the UK and other countries in accordance with clinical guidelines^14–16^. Our results suggest its use could avoid the more time-consuming and costly requirement for pathologist review in more than three quarters of cases. Indeed, the demonstration that the randomly selected subset of 198 tumours with ≥20% epithelial positivity for all MMR proteins were all confirmed as MMRp enabled us to avoid pathologist review for the remainder of this group, totalling roughly two-thirds of tumours in the TransSCOT cohort. Furthermore, AIMMeR was able to identify tumours with combined MLH1-PMS2 loss with high accuracy, and could potentially be combined with reflex *BRAF* mutation or *MLH1* promoter methylation analysis to streamline testing for Lynch syndrome^20,21^. Identification of single protein loss by AIMMeR proved more challenging, and further work is required to improve this to a level suitable for clinical application.

While the prognostic value of MMRd in CRC has been demonstrated previously in trial samples^6,7^, our study confirms that this holds in patients treated with oxaliplatin^9,30^. It is also the first study to show that MMRd tumours have similar outcomes whether treated with 3 or 6 months of such treatment, although our study was only powered to detect a large difference between these groups (see Methods). While the statistically significant interaction we found between MMRd and chemotherapy regimen (CAPOX vs FOLFOX) is intriguing, this must be interpreted cautiously. Choice of chemotherapy regimen in the SCOT trial was not randomised but rather at the discretion of the treating physician, and the similar recurrence rates between MMRp and MMRd patients treated with FOLFOX is discordant with previous results^30^. Firm conclusions regarding the relationship between MMR status and duration and type of adjuvant chemotherapy in CRC await results of adequately powered pooled analyses of multiple studies.

Strengths of our study include its large size, relatively homogeneous patient cohort, high-quality curated demographic, pathological and outcome data, and multiple recruiting sites. The latter is especially relevant, as the performance of AI methods for clinical image analysis often varies between single centres, possibly owing to different protocols for sample fixation, processing and other factors^31^. Our study also has limitations. For logistical reasons, we used images from MMR immunostaining of TMA cores rather than whole slides, and it will be important to evaluate performance in these, and in diagnostic biopsies as neoadjuvant approaches become more widespread^12,13,32^. The absence of data on *BRAF* mutation or *MLH1* promoter methylation and of germline or somatic MMR gene mutations meant we were unable to determine the cause of single MMR protein loss in cases where this was found. Finally, the lack of immunophenotyping of these samples precluded detailed correlation of MMR status with immune infiltrate – analysis of which could inform future immunotherapy studies^33^.

In conclusion, we developed AIMMeR – a single-cell method to identify MMRd in CRC high accuracy and, following pathologist review in a representative subset of cases, confirm its prognostic and predictive value in the SCOT trial cohort. Our study extends the potential applications of AI in CRC diagnostic pathology, and holds promise for clinical implementation.

## METHODS

### Patient selection for biomarker study

Details of the SCOT trial (ISRCTN59757862) have been reported previously^27^. In summary, the study compared the efficacy of 12 vs 24 weeks of oxaliplatin-based adjuvant chemotherapy following curative-intent resection of stage III or high-risk stage II (any of: pT4 primary, tumour obstruction, <10 lymph nodes harvested, grade 3 histology, perineural/extramural venous/lymphatic invasion) colorectal cancer. 6,088 patients were randomised across 237 sites between March 2008 and November 2013. The study met its primary endpoint, with the shorter course of chemotherapy confirmed to be non-inferior (HR=1.01, 95% CI= 0.91–1·11, test for non-inferiority *P*=0·012)^27^ and associated with improved quality of life. Following informed consent, 3,076 patients from 142 recruiting centres donated tissue and blood samples for research as part of the TransSCOT sub-study. Demographic and clinicopathological characteristics of the TransSCOT population were similar to those of the study population as a whole (data not shown).

### Tissue microarrays and immunohistochemistry

Tissue microarrays (TMAs) were constructed from 0.6mm punched cores from formalin-fixed paraffin embedded (FFPE) blocks following review by the TransSCOT pathologists (KO and NM). 2,352 cases with adequate tumour content (epithelial cell fraction of greater than 30%) were included in TMAs. 1,788 of these cases had two cores taken from the tumour centre (TC) and two from the invasive margin (IM) (i.e. a total of four cores per case), while 564 cases had sufficient tumour for additional replicate cores to total of eight cores per case (four CT, four IM). Immunohistochemistry (IHC) for MMR proteins MLH1, MSH2, MSH6 and PMS2 was performed in an accredited UK National Health Service (NHS) diagnostic pathology laboratory (Queen Elizabeth University Hospital, NHS Greater Glasgow & Clyde Trust, Glasgow, UK) to clinical standards by standard methods using ISO approved antibodies and concentrations: MLH1 clone ES05, Leica (Newcastle, UK), Lot no. 6063898, (1:100); MSH2 clone 79H11, Leica, Lot no. 72212, (undiluted); MSH6 clone EP49, Dako (Glostrup, Denmark), Cat no. 1164717, (1:80); PMS2 clone EP51, Dako– Cat no. 1160500, (1:50). Stained slides were scanned using a Hamamatsu NanoZoomer (Hamamatsu, Welwyn Garden City, UK) scanner at 40x and a resolution of 0.22 micron per pixel.

### Artificial Intelligence-based single cell classification and mismatch repair protein quantification

Expression of MMR proteins was quantified on the digitial TMA slides at the single nuclei level using HALO digital image analysis software version 3.3 (Indica Labs, Corrales, NM, USA). First, TMAs were segmented into spots corresponding to individual cores and subjected to rigorous visual quality control (QC). Spots with missing cores, damaged or insufficient tumour tissue were excluded from further analysis. For nuclear segmentation, the HALO AI nuclear Nuclei Seg tool are pre-trained to segment H&E- and DAB-stained nuclei on brightfield images. Additional examples of nuclei (n=2761, total area=825µm^2^) and background (n=352, total area=1698µm^2^) were manually annotated and added to the training set to achieve optimal nuclear segmentation results in immunohistochemically stained tissue. Accuracy of nuclear segmentation was verified by visual pathologist review. The HALO AI Nuclei Phenotyper algorithm was then trained to classify segmented nuclei or other objects into one of the following classes according to cell or object type and individual MMR protein expression (derived from DAB staining): (i) positive tumour cells; (ii) negative tumour cells; (iii) positive stromal cells; (iv) negative stromal cells; (v) lymphocytes (positive and negative); (vi) strong background (e.g., nuclei in out-of-focus cores or strongly DAB-stained non-nuclear objects such as tissue folds, apoptotic bodies, and cellular debris); or (vii) weak background (e.g. weakly DAB-stained objects, such as mucus or extracellular matrix components) (Supplementary Figure S1A). Initial training of the Nuclei Phenotyper used approximately 21,000 annotated nuclei each from all five nuclear classes and background class (strong and weak background, cumulatively) for a total of (almost 127 000 annotated nuclei (Supplementary Table S1). A held-out test set of ∼9000 annotated nuclei was used to measure the accuracy of the Nuclei Phenotyper (Supplementary Table S1). Mark-up images for nucleus segmentation and classification were generated, and the accuracy of nuclear classification was confirmed on the held-out test set and by pathologist review (Supplementary Table S1, Supplementary Figure S1B). The final combined method – AIMMeR – was used to estimate the number of cells or objects in each of the seven classes above for each of the four MMR proteins in all 5,832 TMA cores from all 2,352 cases.

### Pathologist review of cases

Images of MMR staining from cases for pathological review were independently reviewed by two expert GI pathologists (VK and AE) blinded to the results of AIMMeR assessment and those of each other. For each case, pathologists classified MMR status as retained or lost, and recorded the exact combination of proteins lost in the latter case. Cases with failed staining were documented as such. Pathologists also noted any unusual patterns of staining, and the presence of artefacts which could conceivably impact the performance of automated analysis. The results of individual pathologist review were then combined with each other and with the AIMMeR results, and discordances between individual pathologists and between pathologist and AIMMeR noted. Consensus pathologist ground truth was then established as follows:

i. cases where the combination of MMR protein expression was fully concordant across both pathologists and AIMMeR (e.g. classified as retained by all, or classified as combined MLH1-PMS2 loss by all), were documented as per the unanimous classification
ii. cases where the combination of MMR protein expression was discordant, either between pathologists, of between pathologists and AIMMeR results were discussed at a discrepancy meeting. At this, images were reviewed and consensus on the final ground truth classification was reached by discussion between pathologists, and the putative reason for discordance was recorded.
iii. for a subset of 84 cases consensus ground truth was taken following dual pathologist review at a discrepancy meeting without individual pathologist review beforehand. These cases were not used for determination of inter-rater reliability metrics between individual pathologists and AIMMeR results.

AIMMeR, individual pathologist and consensus pathologist classification were then used to establish performance of AIMMeR compared with ground truth and individual pathologist review.

### Data processing and analysis of AIMMeR performance

AIMMeR-derived core level data and image/case metadata were stored as a csv file and processed to obtain case-level results and summary data. Correlation between markers was evaluated by parametric Pearson r and non-parametric Spearman ρ. AIMMeR performance for detection of MMRd was determined by calculation of area under the receiver-operator curve (AUROC) and Youden index, using the consensus pathologist review as ground truth. 95% confidence intervals and out of bag estimates were obtained by bootstrap (n=1,000). Inter-rater reliability ratings were calculated using Cohen’s Kappa and Gwet AC1 for cases in which pathologist review was done. As both of these metrics are influenced by the prevalence of groups for classification, and our selection of cases for review was biased towards cases with MMRd, we calculated predicted values for analysis of the whole cohort, based on the assumption that all cases with ≥20% positive epithelial cells by AIMMeR would be confirmed as MMRd by pathologists (as we determined was the case for the 198 selected at random). The relationship between classification of MMR status and the combination of MMR protein expression determined by AIMMeR and by consensus pathologist review was illustrated by Sankey plots.

### Biomarker and statistical analyses

Associations between MMR status and clinicopathological characteristics of SCOT trial patients were determined by parametric t test and by Chi-square test in the case of continuous and categorical data respectively. Prognostic and predictive analyses of MMR loss were performed and reported in accordance with the REMARK guidelines^34^, as detailed in Table S8. The endpoint for time-to-event analyses was recurrence-free interval (RFI) of CRC, defined as the time from randomization to CRC relapse, with censoring at last contact or death in case of no recurrence. Survival curves were plotted using the Kaplan-Meier method and compared by the log-rank test. Hazard ratios (HRs) were determined by univariable analysis, and by multivariable analysis adjusted for confounders using Cox proportional hazards models. Covariables for inclusion in multivariable models were prespecified, and no variable selection was performed. Inspection of scaled Schoenfeld residuals revealed violation of proportional hazards for analysis of MMR status owing to early recurrences in the MMRd group; hazard ratios should thus be interpreted in the light of this but are preferred over alternatives such as restricted mean survival time (RMST)^35^ for clinical interpretability and consistency with existing literature^36^. Time to event analyses used all informative cases and excluded cases with missing data (i.e no imputation was performed). The sample size was not pre-determined. A power calculation was performed based on 2,000 cases with 500 recurrences (ie similar frequency to the total trial population), assuming prevalence of MMRd or 0.1 and equal proportions of patients treated with 3 months and 6 months chemotherapy in the MMRd and MMRp groups. This demonstrated power to detect an MMRd*chemotherapy duration interaction with difference in hazard ratios of 2.3 or greater, using a 1-λ of 0.8 and a two-sided αof 0.05. Sample sizes and methods used for statistical analyses are provided in the text and figure legends where reported. All statistical tests were two-sided, and hypothesis testing was performed at the 5% significance level. All analyses were performed using R (Comprehensive R Network) version 4.2.2 (2022-10-31) using R Studio version RStudio 2022.07.1, build 554. Plots were exported as vector graphics. Scanned slide images were resized and cropped in Photoshop (Adobe, San Jose CA, USA). Images and figure panels were edited in Illustrator (Adobe). R packages used in this study included: Tidyverse version 1.3.2; ggplot2 version 3.4.0; ggpubr version 0.4.0; cowplot version 1.1.1; stringr version 1.4.1; riverplot version 0.1.0; Tidyverse version 1.3.2; corrplot version 0.9.2; ggcorrplot version 0.1.4; irr version 0.84.1; irrCAC version 1.0; survminer version 0.4.9.oc

## Data Availability

The datasets pertaining to the SCOT trial used during the current study are available from the TransSCOT collaboration on reasonable request. Applications for analysis of TransSCOT samples are welcome and should be addressed to JH

## ADDITIONAL INFORMATION

### TransSCOT group

The TransSCOT Trial Management Group includes (alphabetical order): David Church^1^, Enric Domingo^2^, Joanne Edwards^3^, Bengt Glimelius^4^, Ismail Gogenur^5^, Andrea Harkin^6^, Jen Hay^7^, Timothy Iveson^8^, Emma Jaeger^2^, Caroline Kelly^6^, Rachel Kerr^2^, Noori Maka^7^, Hannah Morgan^7^, Karin Oien^7^, Clare Orange^9^, Claire Palles^10^, Campbell Roxburgh^3^, Owen Sansom^11^, Mark Saunders^12^, Ian Tomlinson^2^.

^1^Cancer Genomics and Immunology Group, The Wellcome Centre for Human Genetics, University of Oxford UK; ^2^Department of Oncology, University of Oxford, UK; ^3^School of Cancer Sciences, University of Glasgow, Glasgow, UK; ^4^Uppsala University, Uppsala, Sweden; ^5^Centre for Surgical Science, Zealand University Hospital, Denmark; ^6^CRUK Glasgow Clinical Trials Unit, University of Glasgow, Glasgow, UK; ^7^Glasgow Tissue Research Facility, University of Glasgow, Queen Elizabeth University Hospital, Glasgow, UK; ^8^University of Southampton, Southampton, UK; ^9^NHS Greater Glasgow and Clyde Biorepository, Glasgow, UK; ^10^University of Birmingham, Birmingham, UK; ^11^CRUK Beatson Institute of Cancer Research, Garscube Estate, Glasgow, UK; ^12^The Christie NHS Foundation Trust, Manchester, UK

## Acknowledgements

We would like to thank the patients who participated in the SCOT trial and consented for their samples to be used for correlative research, as well as the recruiting clinicians and study team. We are also grateful to NHSGGC for performing immunostaining and GTRF (Glasgow University) for TMA construction and scanning.

## Ethical approval and consent to participate

Ethical approval for patient recruitment and sample collection in the SCOT trial was approved centrally and at all recruiting centres. Ethical approval for anonymized tumour molecular analysis was granted by Oxfordshire Research Ethics Committee B (Approval No 05\Q1605\66).

## Author Contributions

Conceptualization: MN, DNC, VHK

Data curation: MN, FJ, AKR, LG, TT, AH, TI, MS, RK, KO, NM, JH

Formal Analysis: MN, FJ, LG, TT, AE, ED, DNC, VHK

Funding acquisition: IT, OS, VHK, DNC

Investigation: MN, FJ, DNC, VHK

Methodology: MN, VHK, DNC

Project administration: JH, VHK, DNC

Resources: AKR, AH, TI, MS, RK, KO, NM, JH, JE, IT, OS, CK, TransSCOT group, DNC

Software: MN, VHK

Supervision: VHK, DNC

Validation: MN, VHK, DNC

Visualization: MN, FJ, ED, VHK, DNC

Writing – original draft: DNC

Writing – review & editing: All authors

## Prognostic and predictive value of single cell, AI-based detection of DNA mismatch repair deficiency in colorectal cancer

### Supplementary material

**Table S1.**
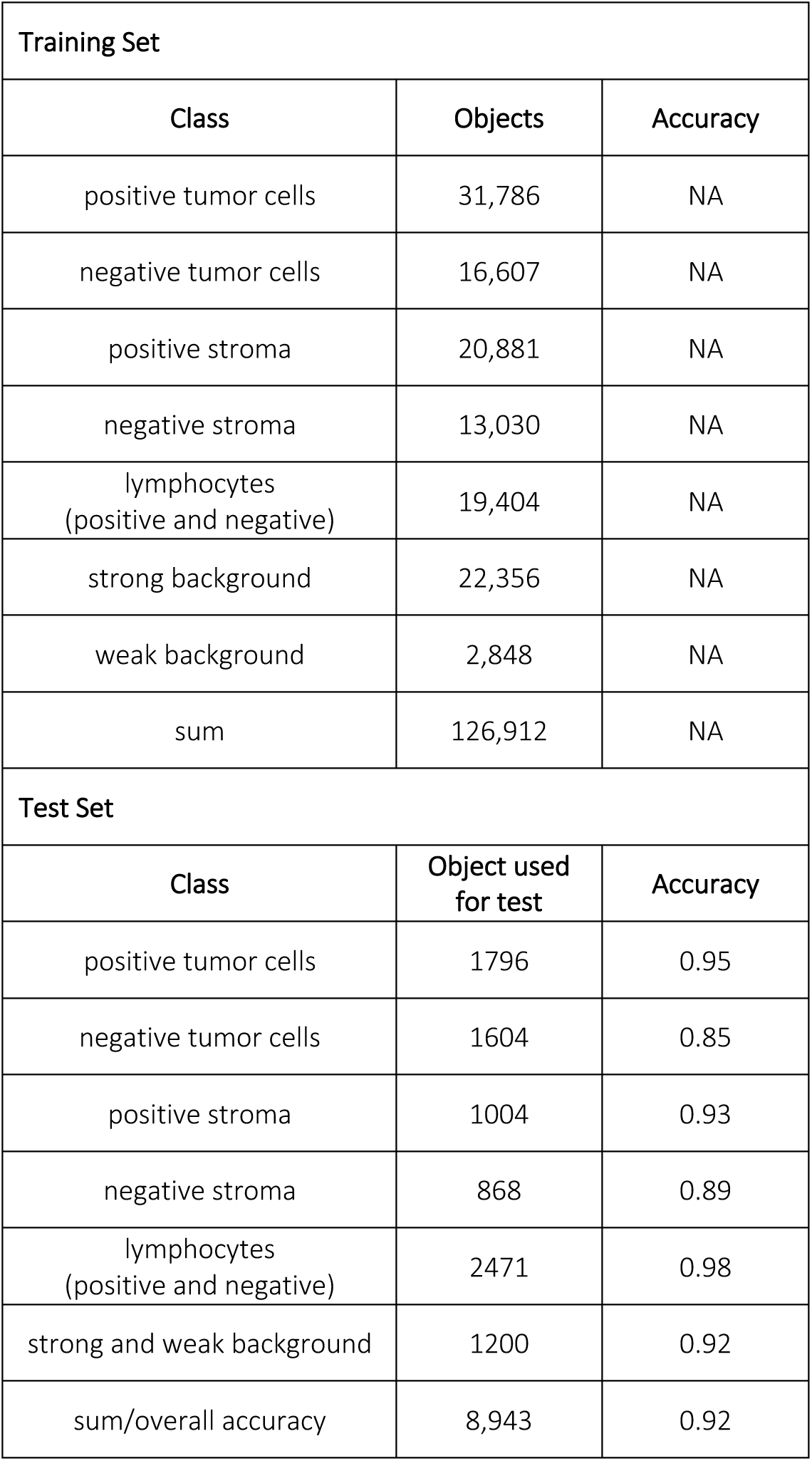
Details of training and test set used for the AIMMeR development in HALO digital image analysis software v3.3.

| Training Set |  |  |
| --- | --- | --- |
| Class | Objects | Accuracy |
| positive tumor cells | 31,786 | NA |
| negative tumor cells | 16,607 | NA |
| positive stroma | 20,881 | NA |
| negative stroma | 13,030 | NA |
| lymphocytes<br>(positive and negative) | 19,404 | NA |
| strong background | 22,356 | NA |
| weak background | 2,848 | NA |
| sum | 126,912 | NA |
| Test Set |  |  |
| Class | Object used<br>for test | Accuracy |
| positive tumor cells | 1796 | 0.95 |
| negative tumor cells | 1604 | 0.85 |
| positive stroma | 1004 | 0.93 |
| negative stroma | 868 | 0.89 |
| lymphocytes<br>(positive and negative) | 2471 | 0.98 |
| strong and weak background | 1200 | 0.92 |
| sum/overall accuracy | 8,943 | 0.92 |

**Table S2.** Agreement between AI and pathologist review for classification of MMR status and protein loss. Individual pathologist review of MMR status and individual MMR protein expression was performed blinded to results of AI-based analysis and the interpretation of the other pathologist. All cases with discordance between AI and one or both pathologists, as well as all cases where individual pathologists were discordant were reviewed at a discrepancy meeting, with final status resolved by discussion. *excludes subset of cases with consensus pathology review but not blinded individual pathological review. †Predicted values are calculated based on the assumption of equivalent concordance between AI and pathologist review in all 1,529 cases with ≥20% epithelial cells positive for all MMR proteins as that obtained by comparison of the randomly-selected subset of 198 cases (100% agreement).

| Group | Comparison | Number of cases | Cohen kappa (95% CI) | Gwet AC1 (95% CI) |
| --- | --- | --- | --- | --- |
| Cases with consensus pathologist review ± blinded individual pathologist review |  |  |  |  |
| MMR status (retained vs lost) | AI vs pathologist consensus | 685 | 0.79 (0.75–0.84) | 0.87 (0.84–0.90) |
|  | AI vs pathologist 1 (AE) | 601* | 0.82 (0.78–0.87) | 0.89 (0.86–0.92) |
|  | AI vs pathologist 2 (VK) | 601* | 0.79 (0.74–0.84) | 0.85 (0.82–0.89) |
|  | Pathologist 1 vs 2 | 601* | 0.88 (0.84–0.92) | 0.92 (0.89–0.94) |
| Individual MMR protein loss | AI vs pathologist consensus | 685 | 0.69 (0.65–0.73) | 0.82 (0.79–0.85) |
|  | AI vs pathologist 1 (AE) | 601* | 0.66 (0.62–0.71) | 0.79 (0.76–0.83) |
|  | AI vs pathologist 2 (AE) | 601* | 0.66 (0.62–0.71) | 0.79 (0.75–0.82) |
|  | Pathologist 1 vs 2 | 601* | 0.84 (0.81–0.88) | 0.91 (0.88–0.93) |
| Predicted values† in total study population |  |  |  |  |
| MMR status (retained vs lost) | AI vs pathologist consensus | 685 analysed<br>1,331 predicted† | 0.85 (0.82–0.88) | 0.96 (0.95–0.97) |
| Individual MMR protein loss | AI vs pathologist consensus | 601 analysed<br>1,331 predicted† | 0.75 (0.71–0.79) | 0.94 (0.93–0.95) |

**Table S3.** Classification of MMR protein loss by AI and consensus pathology review by type of MMR loss.

| AI classification |  | Consensus pathologist classification |  | PPV for MMR status | PPV for protein loss |
| --- | --- | --- | --- | --- | --- |
| Group | Number | Group | Number |  |  |
| MLH1 & PMS2 or MSH2 & MSH6 loss | 147 | MLH1/PMS2 or MSH2/MSH6 | 140 | 0.966 | 0.952* |
|  |  | single protein | 2 |  |  |
|  |  | Other loss | 0 |  |  |
|  |  | retained | 2 |  |  |
|  |  | fail | 3 |  |  |
| Single MMR protein loss | 89 | MLH1/PMS2 or MSH2/MSH6 | 35 | 0.618 | 0.213† |
|  |  | single protein | 19 |  |  |
|  |  | other loss | 1 |  |  |
|  |  | retained | 22 |  |  |
|  |  | fail | 12 |  |  |
| Other MMR loss | 26 | MLH1/PMS2 or MSH2/MSH6 | 11 | 0.577 | 0.115‡ |
|  |  | single protein | 1 |  |  |
|  |  | other loss | 3 |  |  |
|  |  | retained | 1 |  |  |
|  |  | fail | 10 |  |  |
| MMRp | 423 | MLH1/PMS2 or MSH2/MSH6 | 14 | 0.955 | 0.955§ |
|  |  | single protein | 3 |  |  |
|  |  | Other loss | 0 |  |  |
|  |  | retained | 404 |  |  |
|  |  | fail | 2 |  |  |

\* denotes PPV for combined MLH1 & PMS2 loss or combined MSH2 & MSH6 loss; † denotes PPV for single MMR protein loss; ‡ denotes PPV for other patterns of MMR protein loss; § denotes PPV for retained MMR expression

**Table S4.** Reasons for discordance between AI and consensus pathologist classification.

| Reason for discordance | MMR status discordant |  | MMR status concordant but protein loss discordant |  |
| --- | --- | --- | --- | --- |
|  | N | % | N | % |
| Immunostaining failed | 27 | 39.1 | NA | NA |
| Immunostaining weak/heterogenous | 34 | 49.3 | 35 | 70.0 |
| Background/cytoplasmic immunostaining | 1 | 1.4 | 2 | 4.0 |
| Technical: tissue folded | 1 | 1.4 | 0 | 0.0 |
| Technical: shadow/out of focus | 2 | 2.9 | 4 | 8.0 |
| Atypical epithelial morphology | 2 | 2.9 | 2 | 4.0 |
| Subclonal alteration | 1 | 1.4 | 5 | 10.0 |
| No reason identified | 1 | 1.4 | 2 | 4.0 |

**Table S5.** Reasons for discordance between individual pathologist classification.

| Reason for discordance | MMR status discordant |  | MMR status concordant but protein loss discordant |  |
| --- | --- | --- | --- | --- |
|  | N | % | N | % |
| Immunostaining failed | 18 | 47.4 | 0 | NA |
| Immunostaining weak/heterogenous | 11 | 28.9 | 8 | 50 |
| Background/cytoplasmic immunostaining | 0 | 0 | 1 | 6.3 |
| Technical: tissue folded | 1 | 2.6 | 0 | 0 |
| Technical: shadow/out of focus | 3 | 7.9 | 0 | 0 |
| Technical: other | 2 | 5.3 | 1 | 6.3 |
| Atypical epithelial morphology | 1 | 2.6 | 0 | 0 |
| Subclonal alteration | 2 | 5.3 | 0 | 0 |
| No reason identified | 0 | 0 | 6 | 37.5 |

**Table S6.** Clinicopathological characteristics of SCOT trial by MMR status.

|  | MMRp |  | MMRd |  | p-value<sup>2</sup> |
| --- | --- | --- | --- | --- | --- |
| Total | 1,759 |  | 229 |  |  |
| Age |  |  |  |  |  |
| Median IQR |  |  |  |  | <0.001 |
| <70 | 1,302 | 74 | 138 | 60 |  |
| >/70 | 457 | 26 | 91 | 40 |  |
| Sex |  |  |  |  | <0.001 |
| Female | 669 | 38 | 133 | 58 |  |
| Male | 1,090 | 62 | 96 | 42 |  |
| Performance status |  |  |  |  |  |
| 0-1 | 1,759 | 100 | 229 | 100 | 1.0 |
| ≥2 | 0 | 0 | 0 | 0 |  |
| pT stage |  |  |  |  | <0.001 |
| 1-2 | 137 | 7.8 | 3 | 1.3 |  |
| 3 | 1,042 | 59 | 131 | 57 |  |
| 4 | 541 | 31 | 95 | 41 |  |
| Unknown | 39 | 2.2 | 0 | 0 |  |
| N stage |  |  |  |  | <0.001 |
| 0 | 315 | 18 | 76 | 33 |  |
| 1 | 946 | 54 | 109 | 48 |  |
| 2 | 457 | 26 | 44 | 19 |  |
| NA | 39 | 2.2 | 0 | 0 |  |
| AJCC disease stage |  |  |  |  | <0.001 |
| 2 | 314 | 18 | 76 | 33 |  |
| 3 | 1,402 | 80 | 152 | 66 |  |
| Unknown | 42 | 2.4 | 0 | 0 |  |
| Stage III Risk Status |  |  |  |  | <0.001 |
| High | 690 | 39 | 76 | 33 |  |
| Low | 712 | 40 | 76 | 33 |  |
| Not applicable (stage II) | 357 | 20 | 77 | 33 |  |
| Tumour Location |  |  |  |  | <0.001 |
| Left | 1,144 | 65 | 36 | 16 |  |
| Right | 586 | 33 | 193 | 84 |  |
| Unknown | 29 | 1.6 | 0 | 0 |  |
| Treatment Regimen |  |  |  |  | 0.5 |
| CAPOX | 1,241 | 71 | 157 | 69 |  |
| FOLFOX | 518 | 29 | 72 | 31 |  |
| Treatment duration |  |  |  |  | 0.2 |
| 12 weeks | 876 | 50 | 124 | 54 |  |
| 24 weeks | 883 | 50 | 105 | 46 |  |

pT –pathological tumour (T) stage; MMR – DNA mismatch repair; MMRp – mismatch repair proficient; MMRd – mismatch repair deficient;
\*determined by unpaired Student's t-test.
†determined by Fisher exact test (in cases which marker status was determined).

**Table S7.** Univariable and multivariable analysis of recurrence-free interval in SCOT trial cohort according to clinicopathologic factors and MMR status.

| Characteristic | Univariate |  |  |  |  | Multivariate |  |  |  |  |
| --- | --- | --- | --- | --- | --- | --- | --- | --- | --- | --- |
|  | Cases | Events | HR<sup>1</sup> | 95% CI<sup>1</sup> | P | Cases | Events | HR<sup>1</sup> | 95% CI<sup>1</sup> | P |
| Age | 1,959 | 464 |  |  |  | 1,942 | 461 |  |  |  |
| <70 |  |  | 1.00 | — |  |  |  | 1.00 | — |  |
| >/70 |  |  | 1.03 | 0.84, 1.26 | 0.78 |  |  | 0.97 | 0.79, 1.19 | 0.8 |
| Sex | 1,959 | 464 |  |  |  | 1,942 | 461 |  |  |  |
| F |  |  | 1.00 | — |  |  |  | 1.00 | — |  |
| M |  |  | 0.96 | 0.79, 1.15 | 0.63 |  |  | 0.97 | 0.80, 1.17 | 0.7 |
| Tumour Stage | 1,945 | 463 |  |  |  | 1,942 | 461 |  |  |  |
| 1-2 |  |  | 1.00 | — |  |  |  | 1.00 | — |  |
| 3 |  |  | 2.75 | 1.50, 5.04 | 0.001 |  |  | 2.76 | 1.50, 5.07 | 0.001 |
| 4 |  |  | 5.29 | 2.88, 9.69 | <0.001 |  |  | 5.27 | 2.85, 9.74 | <0.001 |
| Nodal Stage | 1,943 | 461 |  |  |  | 1,942 | 461 |  |  |  |
| 0 |  |  | 1.00 | — |  |  |  | 1.00 | — |  |
| 1 |  |  | 1.41 | 1.05, 1.87 | 0.020 |  |  | 1.74 | 1.29, 2.33 | <0.001 |
| 2 |  |  | 2.83 | 2.11, 3.79 | <0.001 |  |  | 3.06 | 2.28, 4.12 | <0.001 |
| Sidedness | 1,959 | 464 |  |  |  | 1,942 | 461 |  |  |  |
| Left |  |  | 1.00 | — |  |  |  | 1.00 | — |  |
| Right |  |  | 1.41 | 1.17, 1.69 | <0.001 |  |  | 1.30 | 1.07, 1.58 | 0.008 |
| Regimen | 1,959 | 464 |  |  |  | 1,942 | 461 |  |  |  |
| Capox |  |  | 1.00 | — |  |  |  | 1.00 | — |  |
| Folfox |  |  | 1.11 | 0.92, 1.35 | 0.28 |  |  | 1.01 | 0.83, 1.23 | >0.9 |
| Duration | 1,959 | 464 |  |  |  | 1,942 | 461 |  |  |  |
| 12 weeks |  |  | 1.00 | — |  |  |  | 1.00 | — |  |
| 24 weeks |  |  | 0.91 | 0.75, 1.09 | 0.28 |  |  | 0.92 | 0.76, 1.10 | 0.4 |
| MMR status | 1,959 | 464 |  |  |  | 1,942 | 461 |  |  |  |
| MMRp |  |  | 1.00 | — |  |  |  | 1.00 | — |  |
| MMRd |  |  | 0.69 | 0.50, 0.96 | 0.029 |  |  | 0.62 | 0.44, 0.88 | 0.007 |

<sup>1</sup>HR = Hazard Ratio, CI = Confidence Interval

**Table S8.** Biomarker analyses performed and reported in this study.

| Analysis | Population | Methods | Reported |
| --- | --- | --- | --- |
| RFI by MMR status | Stage II/III CRCs from SCOT trial | Log-rank test, univariable and multivariable adjusted HR | Main text, Figure 5, Table S8 |
| RFI by MMR status | Defined subgroups in SCOT trial | Univariable and multivariable adjusted HR | Figure 5 Table S8 |

RFI – recurrence-free interval; OS – overall survival; HR – hazard ratio. \*Full multivariable model included age, sex, location,

**Figure S1.**
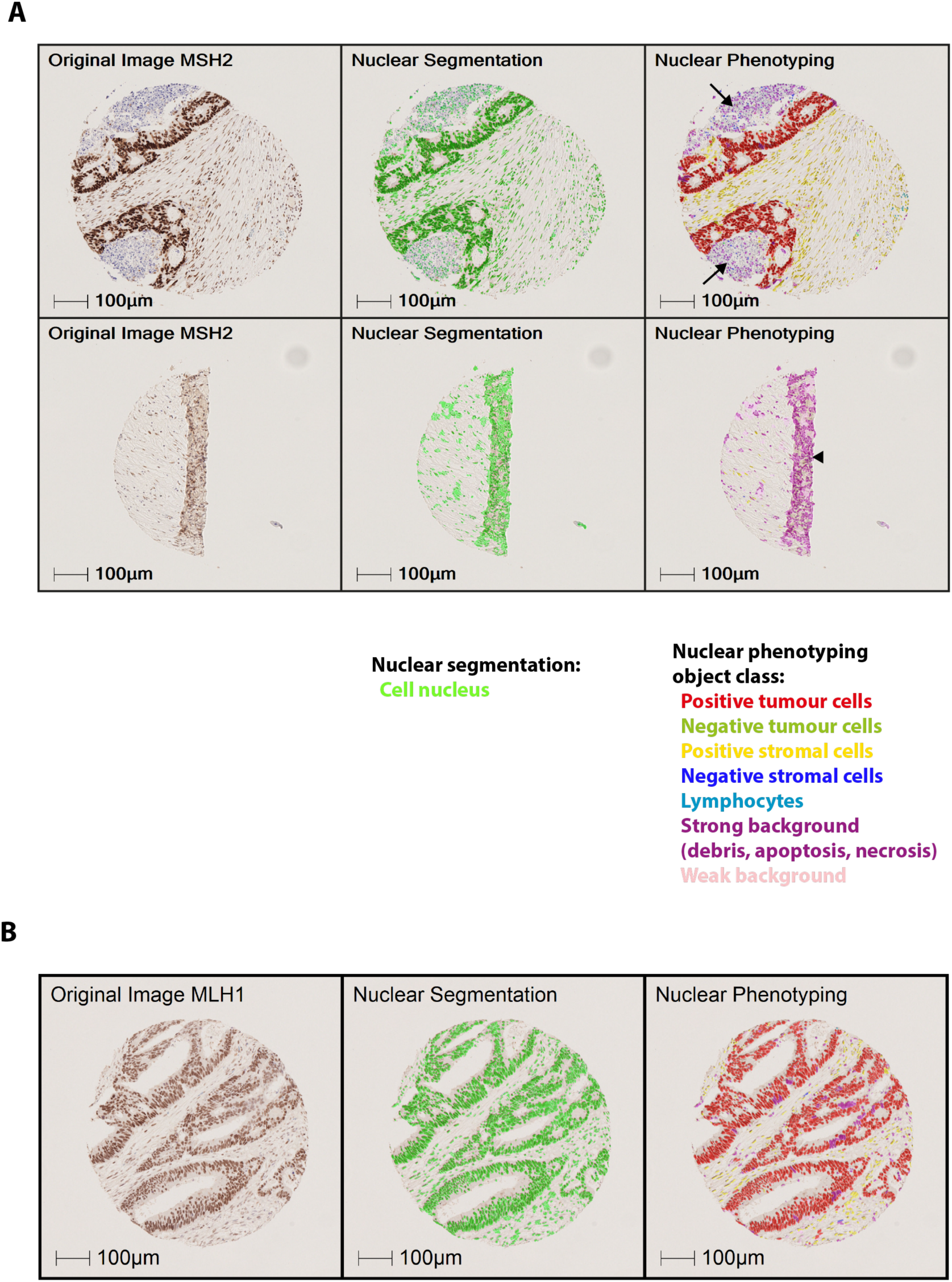
Nuclear segmentation by object class. (A) Illustrative images of AI-based classification of cell nuclei and other objects on tumor sections following IHC for MMR proteins. Panels show the original IHC stained images (left), nuclear segmentation mark-ups (centre) and nuclear phenotyping by object classification (right). Upper panels show the original image (left), nuclear segmentation (center) and classification results (right) of a representative tissue microarray core with tumor cells, stromal cells and intraglandular debris (black arrows) correctly classified. Lower panels show an exemplary core with a folding artefact (black arrow), with the nuclei in the affected area and adjacent out-of-focus tissue regions correctly classified as strong background (uninformative for analysis) by nuclear phenotyping,. Note that partial transparency in nuclear segmentation masks permits visualisation of DAB staining in MSH2 positive cells.

**Figure S2.**
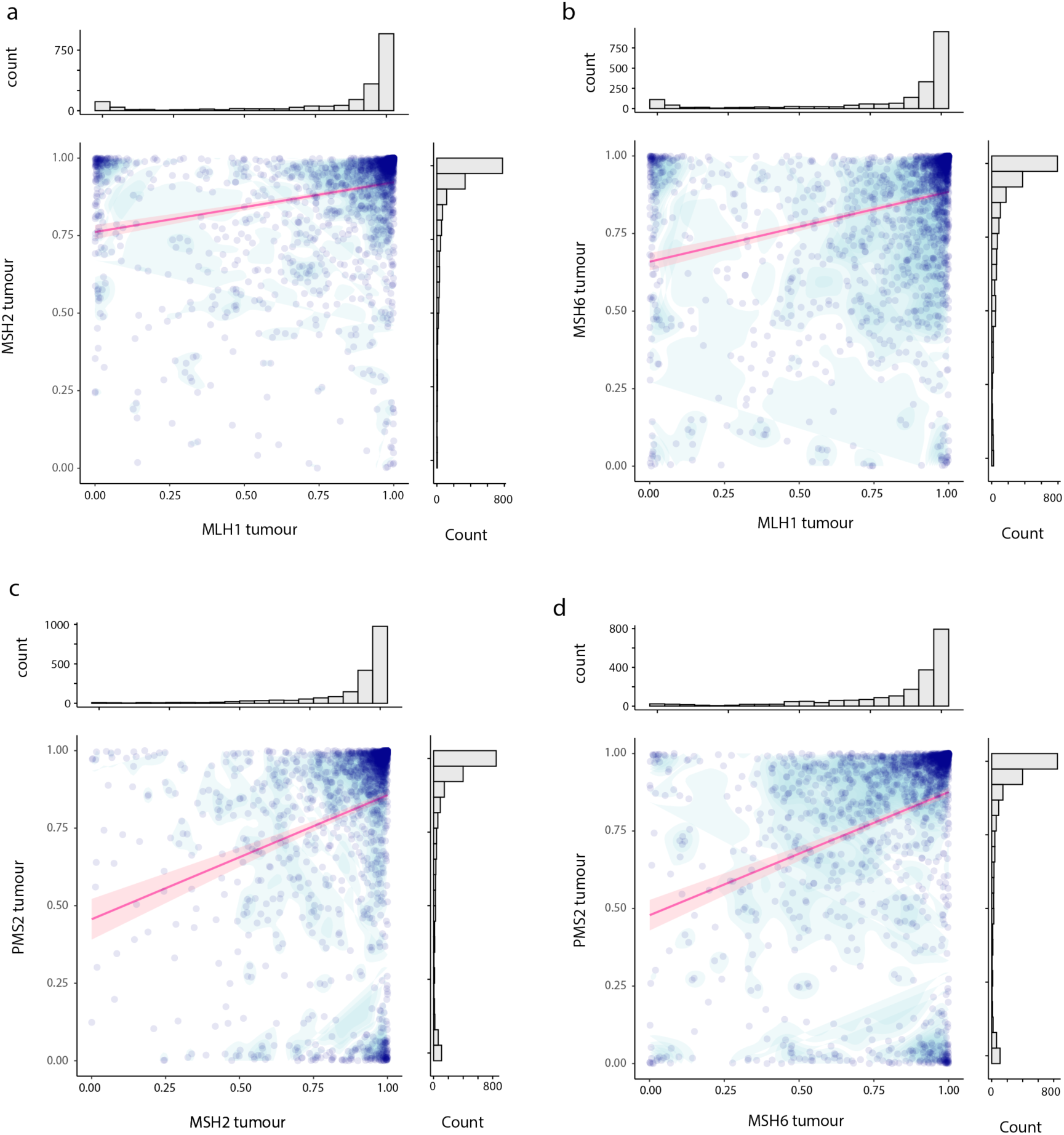
Scatterplots showing correlation between epithelial MMR protein expression across cases. Scatterplots with marginal histograms showing correlation between the proportion of epithelial cells positive for individual MMR proteins for combinations: (a) MLH1 and MSH2; (b) MLH1 and MSH6; (c) MSH2 and PMS2; and (d) MSH6 and PMS2.

**Figure S3.**
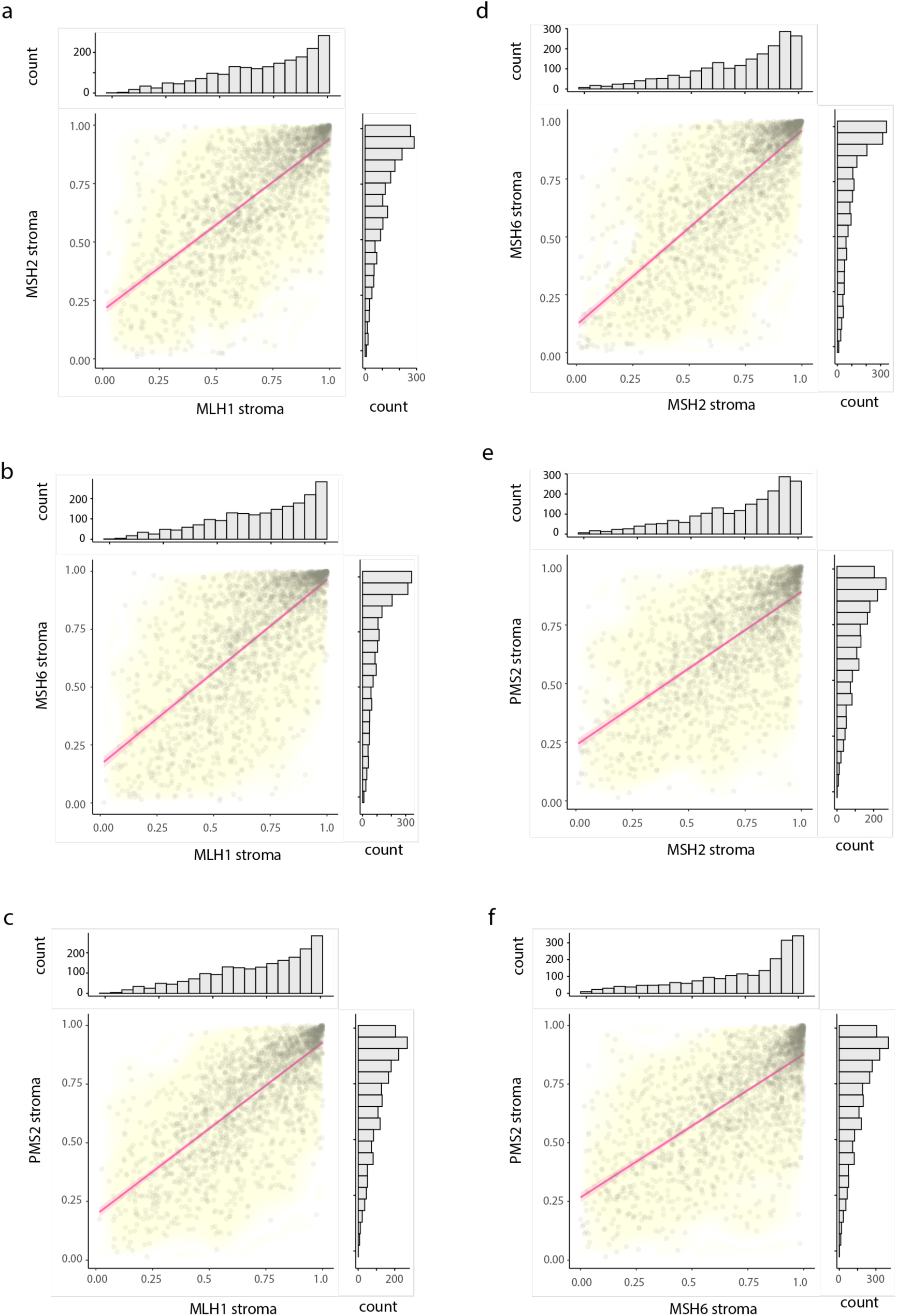
Scatterplots showing correlation between stromal MMR protein expression across cases. Scatterplots with marginal histograms showing correlation between the proportion of stromal cells positive for individual MMR proteins for: (a) MLH1 and MSH2, (b) MLH1 and MSH6, (c) MLH1 and PMS2; (d) MSH2 and MSH6; (e) MSH2 and PMS2 and: (f) MSH6 and PMS2.

**Figure S4.**
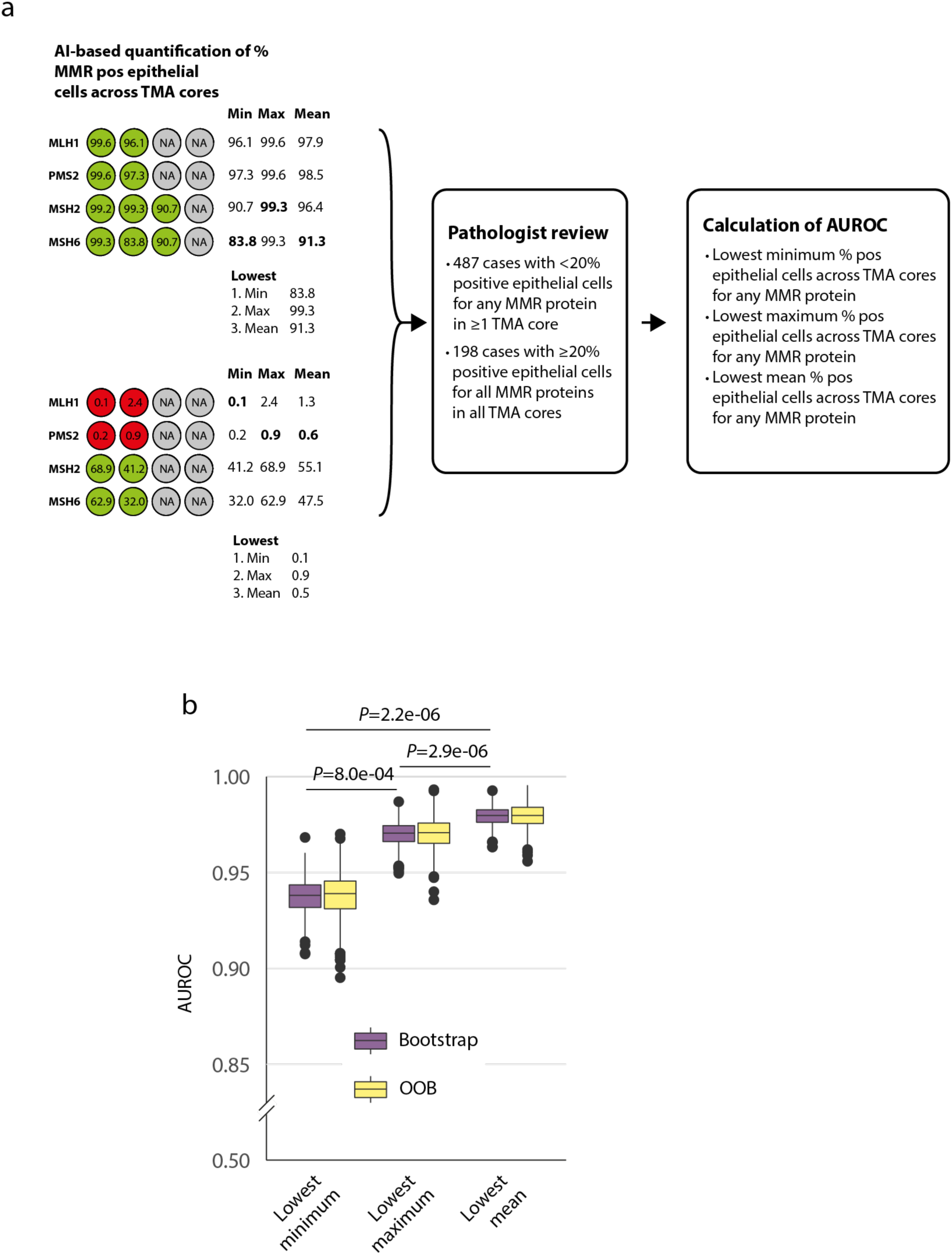
Calculation of AUROC for alternative methods for classification of MMR loss. (a) Schematic showing study workflow and identification of cases for establishing consensus pathologist ground truth for evaluation of AIMMeR performance. (b) AUROC calculated against consensus pathologist ground truth using alternative metrics based on minimum, maximum and mean percentage of cells positive for individual MMR proteins. Boxplots show median, 25^th^ and 75^th^ percentiles ± 1.5 x interquartile range and outlying points obtained from bootstrap with 1000 resamples and corresponding out of bag (OOB estimates). *P* values were obtained by Mann-Whitney U test.

**Figure S5.**
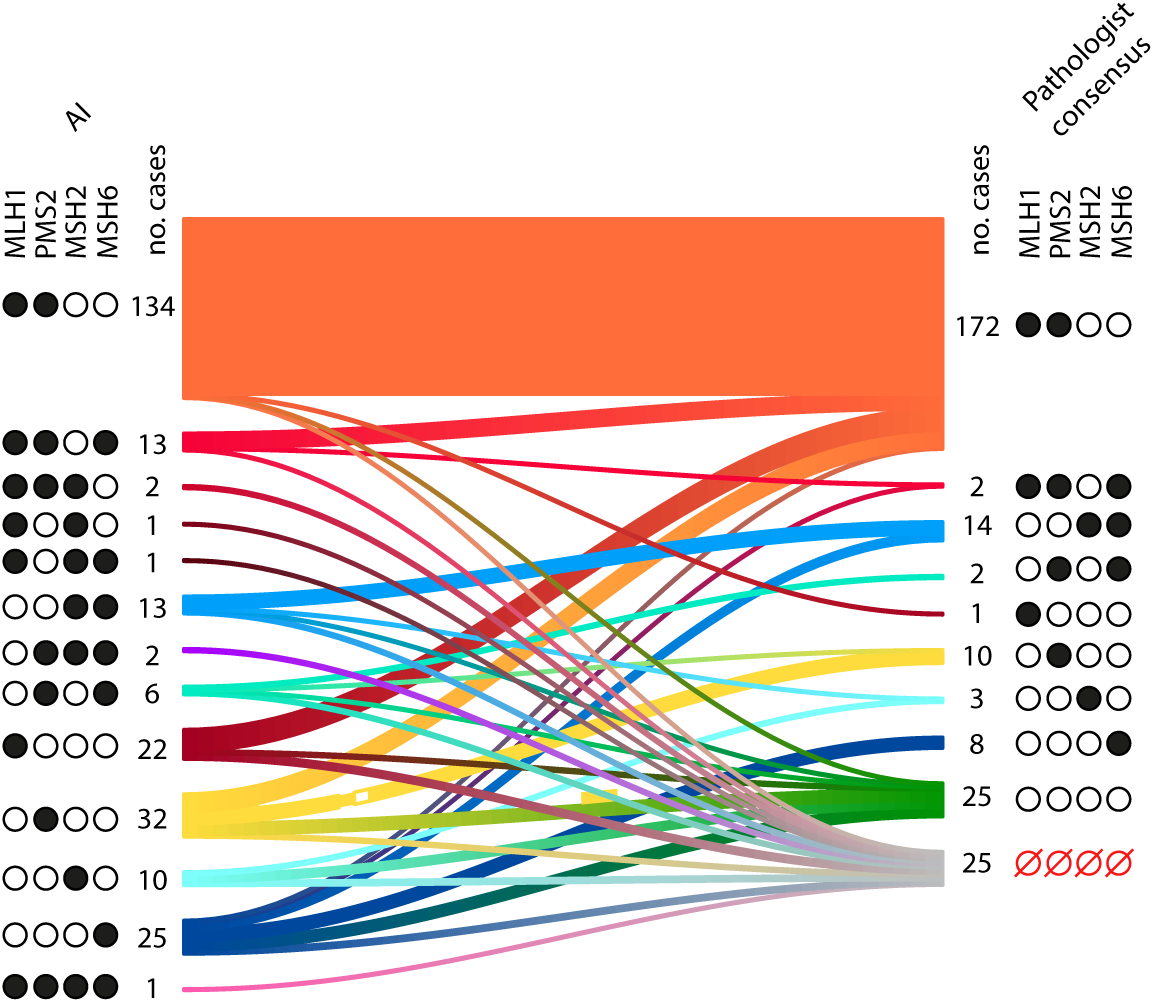
Relationship between AI and consensus pathologist calls for combinations of MMR protein loss. Sankey plot showing flow between initial AI classification of MMR protein loss using cutpoint of 10.7% positive epithelial cells and final consensus pathologist calls. Proteins lost are indicated by closed circles. Open red circles indicate failed cases.

**Figure S6.**
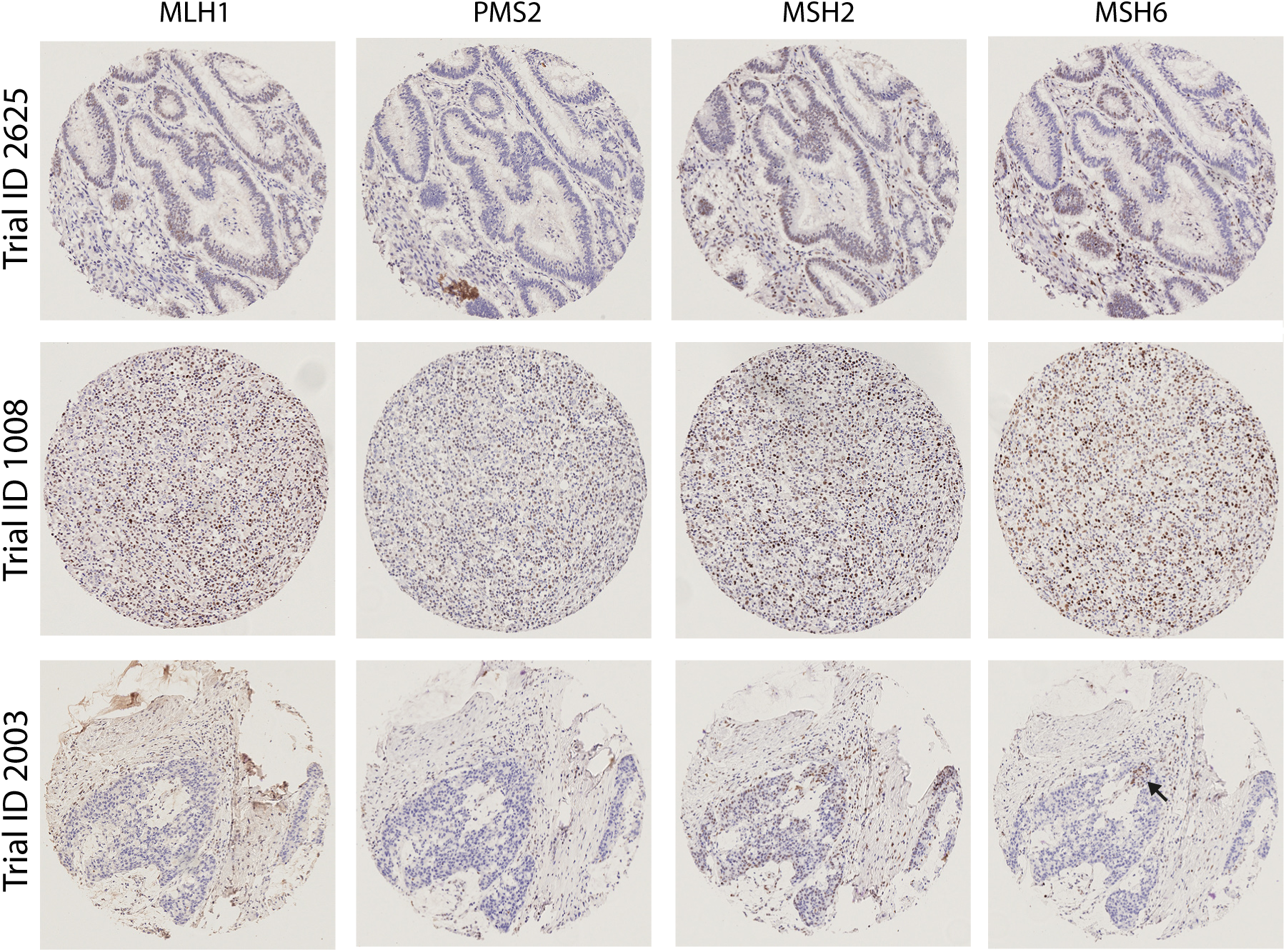
Cases discordant between AI-based and pathologist classification. Representative cases illustrative of causes of AI-pathologist discordance are shown. Upper panel shows failed immunostaining in tumour from participant ID 2625. Immunostain for PMS2 was classified by AI as negative (no positive tumour cells). On pathologist review, this was reclassified to failed staining in view of absence of internal positive controls, and poor quality immunostaining for other MMR proteins. Middle panels show a case misclassified by AI as MMRp as a result of atypical tumour epithelial morphology, in setting of intense lymphocytic infiltrate. On pathologist review, this was reclassified to MMRd with loss of MLH1 and PMS2. Lower panels show a case classified as MMRd by both AI and pathologist review, but discordant for protein loss. AI-based analysis classified this as lacking expression of MLH1, PSM2 and MSH6; the latter owing to MSH6 expression in 2.6% of epithelial cells. Pathologist review identified a small area of retained MSH6 expression (black arrow) in background of loss, leading to reclassification of case as MLH1 and PMS2 deficient with subclonal MSH6 loss.

**Figure S7.**
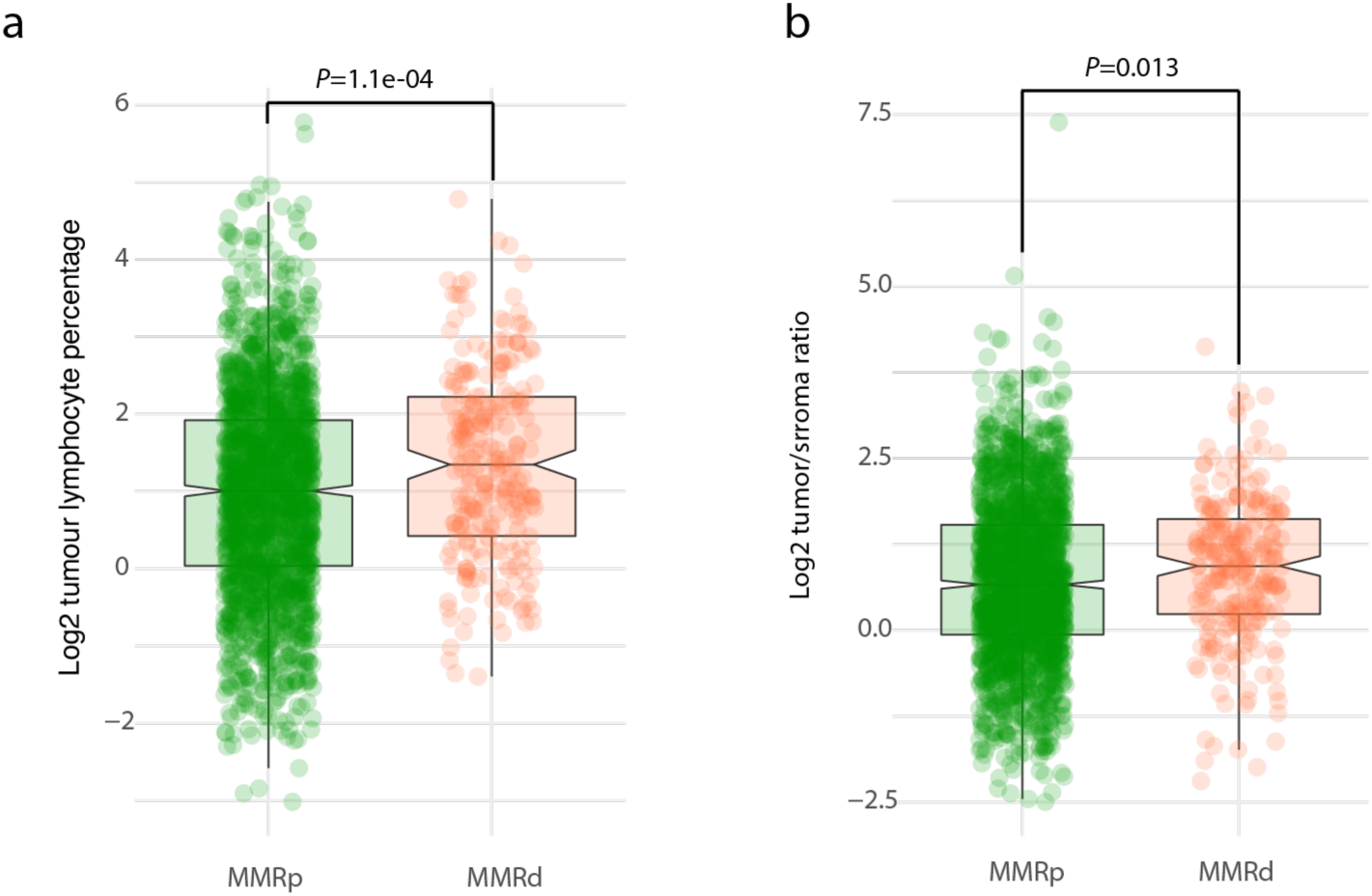
Tumor lymphocytic infiltrate and tumor/stroma ratio by MMR status. (a) Lymphocytes as percentage of all cells within tumor (determined by single-cell AI-based analysis) according to MMR status. (b) Tumor (malignant epithelial) cell/stroma cell ratio (determined by single-cell AI-based analysis) according MMR status.

## REFERENCES

1 Sung, H., et al. Global Cancer Statistics 2020: GLOBOCAN Estimates of Incidence and Mortality Worldwide for 36 Cancers in 185 Countries. CA: a cancer journal for clinicians 71, 209–249, doi:10.3322/caac.21660 (2021).

2 Sinicrope, F. A. Lynch Syndrome-Associated Colorectal Cancer. N Engl J Med 379, 764–773, doi:10.1056/NEJMcp1714533 (2018).

3 Haraldsdottir, S. et al. Colon and Endometrial Cancers with Mismatch Repair Deficiency can Arise from Somatic, Rather Than Germline, Mutations. Gastroenterology, doi:10.1053/j.gastro.2014.08.041 (2014).

4 Mensenkamp, A. R. et al. Somatic mutations in MLH1 and MSH2 are a frequent cause of mismatch-repair deficiency in Lynch syndrome-like tumors. Gastroenterology 146, 643–646.e648, doi:10.1053/j.gastro.2013.12.002 (2014).

5 Kane, M. F. et al. Methylation of the hMLH1 promoter correlates with lack of expression of hMLH1 in sporadic colon tumors and mismatch repair-defective human tumor cell lines. Cancer Res 57, 808–811 (1997).

6 Hutchins, G. et al. Value of mismatch repair, KRAS, and BRAF mutations in predicting recurrence and benefits from chemotherapy in colorectal cancer. Journal of Clinical Oncology 29, 1261–1270, doi:10.1200/JCO.2010.30.1366 (2011).

7 Bertagnolli, M. M. et al. Microsatellite instability and loss of heterozygosity at chromosomal location 18q: prospective evaluation of biomarkers for stages II and III colon cancer--a study of CALGB 9581 and 89803. Journal of Clinical Oncology 29, 3153–3162, doi:10.1200/JCO.2010.33.0092 (2011).

8 Vilar, E. & Gruber, S. B. Microsatellite instability in colorectal cancer—the stable evidence. Nature Reviews Clinical Oncology 7, 153–162, doi:10.1038/nrclinonc.2009.237 (2010).

9 Cohen, R. et al. Microsatellite Instability in Patients With Stage III Colon Cancer Receiving Fluoropyrimidine With or Without Oxaliplatin: An ACCENT Pooled Analysis of 12 Adjuvant Trials. J Clin Oncol 39, 642–651, doi:10.1200/jco.20.01600 (2021).

10 Le, D. T. et al. PD-1 Blockade in Tumors with Mismatch-Repair Deficiency. N Engl J Med 372, 2509–2520, doi:10.1056/NEJMoa1500596 (2015).

11 Overman, M. J. et al. Durable Clinical Benefit With Nivolumab Plus Ipilimumab in DNA Mismatch Repair-Deficient/Microsatellite Instability-High Metastatic Colorectal Cancer. J Clin Oncol 36, 773–779, doi:10.1200/jco.2017.76.9901 (2018).

12 Chalabi, M. et al. Neoadjuvant immunotherapy leads to pathological responses in MMR-proficient and MMR-deficient early-stage colon cancers. Nat Med 26, 566–576, doi:10.1038/s41591-020-0805-8 (2020).

13 Chalabi, M. V., Y.L.; van den Berg, J.; Sikorska, K.; Beets, G.; Lent, A.V.; Grootscholten, M.C.; Aalbers, A.; Buller, N.; Marsman, H.; Hendriks, E.; Burger, P.W.A.; Aukema, T.; Oosterling, S.; Beets-Tan, R.; Schumacher, T.N.; van Leerdam, M.; Voest, E.E.; Haanen, J.B.A.G.. LBA7-Neoadjuvant immune checkpoint inhibition in locally advanced MMR-deficient colon cancer: The NICHE-2 study. Annals of Oncology 33, S808–S869, doi:10.1016/annonc/annonc1089 (2022).

14 Argilés, G. et al. Localised colon cancer: ESMO Clinical Practice Guidelines for diagnosis, treatment and follow-up. Ann Oncol 31, 1291–1305, doi:10.1016/j.annonc.2020.06.022 (2020).

15 Network, N. C. C. Colon Cancer – Version 2.2022, <https://www.nccn.org/professionals/physician_gls/pdf/colon.pdf> (2022).

16 Bartley, A. N. et al. Mismatch Repair and Microsatellite Instability Testing for Immune Checkpoint Inhibitor Therapy: Guideline From the College of American Pathologists in Collaboration With the Association for Molecular Pathology and Fight Colorectal Cancer. Arch Pathol Lab Med 146, 1194–1210, doi:10.5858/arpa.2021-0632-CP (2022).

17 Kather, J. N. et al. Deep learning can predict microsatellite instability directly from histology in gastrointestinal cancer. Nat Med 25, 1054–1056, doi:10.1038/s41591-019-0462-y (2019).

18 Echle, A. et al. Clinical-Grade Detection of Microsatellite Instability in Colorectal Tumors by Deep Learning. Gastroenterology 159, 1406–1416.e1411, doi:10.1053/j.gastro.2020.06.021 (2020).

19 Echle, A. et al. Artificial intelligence for detection of microsatellite instability in colorectal cancer-a multicentric analysis of a pre-screening tool for clinical application. ESMO Open 7, 100400, doi:10.1016/j.esmoop.2022.100400 (2022).

20 Stoffel, E. M. et al. Hereditary colorectal cancer syndromes: American Society of Clinical Oncology Clinical Practice Guideline endorsement of the familial risk-colorectal cancer: European Society for Medical Oncology Clinical Practice Guidelines. J Clin Oncol 33, 209–217, doi:10.1200/jco.2014.58.1322 (2015).

21 Monahan, K. J. et al. Guidelines for the management of hereditary colorectal cancer from the British Society of Gastroenterology (BSG)/Association of Coloproctology of Great Britain and Ireland (ACPGBI)/United Kingdom Cancer Genetics Group (UKCGG). Gut 69, 411–444, doi:10.1136/gutjnl-2019-319915 (2020).

22 Glaire, M., Domingo, E., Nicholson, G., Novelli, M., Lawson, K., Oukrif, D., Kidal, W., Danielsen, H.E., Kerr, R., Kerr, D.J., Tomlinson, I., Church, D.N. Tumour-infiltrating CD8+ lymphocytes as a prognostic marker in colorectal cancer: A retrospective, pooled analysis of the QUASAR2 and VICTOR trials. Journal of Clinical Oncology 36, 3515–3515 (2018).

23 Pagès, F. et al. International validation of the consensus Immunoscore for the classification of colon cancer: a prognostic and accuracy study. Lancet 391, 2128–2139, doi:10.1016/s0140-6736(18)30789-x (2018).

24 Horeweg, N. et al. Prognostic Integrated Image-Based Immune and Molecular Profiling in Early-Stage Endometrial Cancer. Cancer immunology research 8, 1508–1519, doi:10.1158/2326-6066.Cir-20-0149 (2020).

25 Llosa, N. J. et al. The vigorous immune microenvironment of microsatellite instable colon cancer is balanced by multiple counter-inhibitory checkpoints. Cancer Discov 5, 43–51, doi:10.1158/2159-8290.CD-14-0863 (2015).

26 Domingo, E. et al. Somatic POLE proofreading domain mutation, immune response, and prognosis in colorectal cancer: a retrospective, pooled biomarker study. The lancet. Gastroenterology & hepatology 1, 207–216, doi:10.1016/s2468-1253(16)30014-0 (2016).

27 Iveson, T. J. et al. 3 versus 6 months of adjuvant oxaliplatin-fluoropyrimidine combination therapy for colorectal cancer (SCOT): an international, randomised, phase 3, non-inferiority trial. The Lancet. Oncology 19, 562–578, doi:10.1016/s1470-2045(18)30093-7 (2018).

28 Loughrey, M. Q., P; Shepherd, NA. Dataset for histopathological reporting of colorectal cancer, <https://www.rcpath.org/static/c8b61ba0-ae3f-43f1-85ffd3ab9f17cfe6/G049-Dataset-for-histopathological-reporting-of-colorectal-cancer.pdf> (2018).

29 Kather, J. N. et al. Pan-cancer image-based detection of clinically actionable genetic alterations. Nat Cancer 1, 789–799, doi:10.1038/s43018-020-0087-6 (2020).

30 Zaanan, A. et al. Role of Deficient DNA Mismatch Repair Status in Patients With Stage III Colon Cancer Treated With FOLFOX Adjuvant Chemotherapy: A Pooled Analysis From 2 Randomized Clinical Trials. JAMA oncology 4, 379–383, doi:10.1001/jamaoncol.2017.2899 (2018).

31 Van Eycke, Y. R., Allard, J., Salmon, I., Debeir, O. & Decaestecker, C. Image processing in digital pathology: an opportunity to solve inter-batch variability of immunohistochemical staining. Sci Rep 7, 42964, doi:10.1038/srep42964 (2017).

32 Morton, D. et al. Preoperative Chemotherapy for Operable Colon Cancer: Mature Results of an International Randomized Controlled Trial. J Clin Oncol, Jco2200046, doi:10.1200/jco.22.00046 (2023).

33 de Vries, N. L. et al. γδ T cells are effectors of immunotherapy in cancers with HLA class I defects. Nature 613, 743–750, doi:10.1038/s41586-022-05593-1 (2023).

34 McShane, L. M. et al. in Br J Cancer Vol. 93 387–391 (2005).

35 Royston, P. & Parmar, M. K. Augmenting the logrank test in the design of clinical trials in which non-proportional hazards of the treatment effect may be anticipated. BMC Med Res Methodol 16, 16, doi:10.1186/s12874-016-0110-x (2016).

36 Freidlin, B. & Korn, E. L. Methods for Accommodating Nonproportional Hazards in Clinical Trials: Ready for the Primary Analysis? J Clin Oncol 37, 3455–3459, doi:10.1200/jco.19.01681 (2019).

